# “Soluble CD163 changes indicate monocyte association with cognitive deficits in Parkinson’s disease”

**DOI:** 10.1101/2020.05.02.20088500

**Authors:** Sara K. Nissen, Sara A. Ferreira, Marlene C. Nielsen, Claudia Schulte, Kalpana Shrivastava, Dorle Hennig, Anders Etzerodt, Jonas H. Graversen, Daniela Berg, Walter Maetzler, Anne Panhelainen, Holger J. Møller, Kathrin Brockmann, Marina Romero-Ramos

**Affiliations:** DANDRITE & Department of Biomedicine, Aarhus University, Aarhus, Denmark; Department of Clinical Biochemistry, Aarhus University Hospital, Aarhus, Denmark; Center of Neurology, Department of Neurodegeneration and Hertie-Institute for Clinical Brain Research & German Center for Neurodegenerative Diseases, University of Tuebingen, Tuebingen, Germany; Department of Molecular Medicine, University of Southern Denmark, Odense, Denmark; Department of Neurology, Christian-Albrechts University, Kiel, Germany; Institute of Biotechnology, University of Helsinki, Helsinki, Finland

**Keywords:** monocytes, sCD163, biomarkers, cognition, alpha-synuclein

## Abstract

**Background:** Parkinson’s disease (PD) is a neurodegenerative disorder with a significant immune component, as demonstrated by changes in immune biomarkers in patients’ biofluids. However, which specific cells are responsible for those changes is unclear since most immune biomarkers can be produced by various cell types.

**Objectives and methods:** To explore monocyte involvement in PD, we investigated the monocyte-specific biomarker sCD163, the soluble form of the receptor CD163, in cerebrospinal fluid (CSF) and serum in two experiments, and compared it with other biomarkers and clinical data. Potential connections between CD163 and alpha-synuclein were studied *in vitro*.

**Results:** CSF-sCD163 increased in late-stage PD and correlated with the PD biomarkers alpha-synuclein, Tau, and phosphorylated Tau, while it inversely correlated with the patients’ cognitive scores, supporting monocyte involvement in neurodegeneration and cognition in PD. Serum-sCD163 only increased in female patients, suggesting a sex-distinctive monocyte response. CSF-sCD163 also correlated with molecules associated with adaptive and innate immune system activation and with immune cell recruitment to the brain. Serum-sCD163 correlated with pro-inflammatory cytokines and acute phase proteins, suggesting a relation to chronic systemic inflammation. Our *in vitro* study showed that alpha-synuclein activates macrophages and induces shedding of sCD163, which in turn enhances alpha-synuclein uptake by myeloid cells, potentially participating in its clearance.

**Conclusion:** Our data present sCD163 as a potential cognition-related biomarker in PD and suggest a role for monocytes both in peripheral and brain immune responses. This may be directly related to alpha-synuclein’s pro-inflammatory capacity but could also have consequences for alpha-synuclein processing.

## INTRODUCTION

Parkinson’s disease (PD) is characterized by intraneuronal aggregations of alpha-synuclein (α-syn) and dopaminergic neuronal death in the midbrain. PD also involves early and chronic immune activation^1^ where α-syn act as a damage-associated molecular pattern, initiating inflammation and promoting disease^2^. The immune response involves microglia in the brain and blood immune cells such as monocytes/macrophages and lymphocytes. In PD patients, the monocytic population shows increased proliferative and phagocytic capacity, abnormal response to immune stimuli^3, 4^, and an altered transcriptome^5^. PD-derived T cells show a bias towards pro-inflammatory Th1/Th17 phenotypes^6, 7^. Accordingly, the PD immune response involves both innate and adaptive immune components. These peripheral alterations will affect the central nervous system (CNS) since T cells and monocytes infiltrate the brain in PD patients and models^8-10^. This results in changes in cytokines and chemokines in patients’ serum and cerebrospinal fluid (CSF). A recent meta-analysis reports increased IL-1β, TNF-α, IL-6, and IL-10 in PD patients’ blood, whereas in CSF, IL-1β, IL-6, TGF-β1, and CRP are elevated^11^. These biomarkers correlate with PD symptomatology^1^, further supporting their disease relevance.

These PD-related immune biomarkers can be produced by multiple cell types^12^ and cannot truly reveal the unique intervention of each cell population involved in the PD immune response. Conversely, this is not the case with soluble CD163 (sCD163), a well-characterized protein produced exclusively by the monocyte cell lineage, but not by lymphocytes, neurons^13, 14^, or microglia, as confirmed recently by single-cell RNA analysis^15-17^. In the brain, CD163 is expressed only by non-microglia CNS myeloid cells, such as meningeal and perivascular macrophages^17-19^. sCD163 is constitutively produced in serum and CSF^20^ upon immune signals, through mechanisms similar to that releasing TNF-α during inflammatory processes ^21, 22^. Therefore, increased sCD163 levels in biofluids relate to macrophage activation and correlate with the degree of inflammation^14^. In the present study, we analyze sCD163 levels in serum and CSF from patients with early and late PD to evaluate monocytic activation in different PD stages.

## MATERIALS AND METHODS

### Study participants

PD patients were recruited from the outpatient clinic and/or ward for PD, University Hospital of Tuebingen, Germany. Healthy controls (HCs) were assessed as having no neurodegenerative disease. All participants were examined by a neurologist specialized in movement disorders. A PD diagnosis was made according to the UK Brain Bank Society Criteria^23^. Serum and CSF were collected and stored at the Neuro-Biobank, University of Tuebingen and sent for experimental work at Aarhus University, Denmark (see supplement). The study was approved by the local ethics committee (480/2015BO2), with all participants providing informed consent. This cross-sectional study included serum and CSF from idiopathic PD (early [<5y] & late [≥5y from diagnosis]) patients and HC (**Suppl.Fig. 1**). Samples were received and analyzed in two separate experiments with a two-year separation: *Exp#1:* 109 PD patients and 44 HCs (**Suppl.Table1**), and *Exp#2:* 106 PD patients and 16 HCs (**Suppl.Table2)**. Twenty-eight individuals from Exp#1 were also part of Exp#2; however, other aliquots were used. Furthermore, 26 additional serum HC samples remaining from Exp#1 were included in the 40-plex mesoscale analysis in Exp#2 (**Suppl.Table3**).

### Biomarkers measurements

sCD163 was measured in serum and CSF using an in-house ELISA^3, 24^. ELISA was used to measure CSF levels of total human Tau (h-Tau), phosphorylated Threonine 181-Tau (p-Tau), Abeta_1-42_ (Abeta42) (all by Innotest), and total α-syn (Analytica Jena Roboscreen GmbH). MSD MULTI-SPOT Assay System (Mesoscale) was used to measure 40 different molecules in serum and CSF (see supplement).

### α-Synuclein and sCD163 association

Evaluation of sCD163-α-syn binding was studied using Microscale Thermophoresis (MST). Monocyte-derived macrophages (MDMs) from isolated and maturated human monocytes^25^ were stimulated with 100nM, 1µM, or 5µM monomeric or fibrillar α-syn for 6 or 24 hours, followed by sCD163-ELISA measurement in supernatants.

Differentiated THP-1 macrophage-like cells or BV-2 murine microglia cells were used to study ^125^I-α-syn fibril uptake (50 ng/ml) alone or with sCD163 (5µg/ml) as a co-treatment or pre-treatment. Full-length (domain 1-9) or truncated (domain 1-5) sCD163 were used, and when stated, LPS was removed from sCD163 preparations as described^26^ (see supplement).

### Statistical analyses

GraphPad Prism V7, JMP 14, and STATA v15 IC were used for statistical analyses where normality was first informed (see supplement).

## RESULTS

### sCD163 as a biomarker related to cognition (Exp#1)

First, we investigated any PD-related changes in sCD163 in Exp#1: biobank samples from 109 PD patients and 44 HCs (**Suppl.Fig. 1; Suppl.Table1**). CSF-sCD163 was elevated in late PD (**Fig.1A**), with no sex discrepancies. Serum-sCD163 levels were influenced by sex and treatment (two-way ANOVA [F(7, 140)=2.86, p=0.008], sex effect p=0.039, treatment effect p=0.017). Therefore, we separated the data by sex, and to eliminate the treatment variable, we analyzed only treated patients. We found elevated serum-sCD163 levels in females with late PD (vs. early), but not in males (**Fig.1B**). To confirm that changes in CSF-sCD163 were not a simple leakage from serum, we calculated the amount of intrathecally produced and serum-derived sCD163 using the ratio of albumin CSF/serum as reference (when available)^27^. We found no difference in the amount of CSF-sCD163 of serum origin (late PD 0.0132±0.001, early PD 0.0139±0.001). On the other hand, intrathecally produced sCD163 was higher in late PD (0.0739±0.037) than in early PD (0.0601±0.019, p=0.03), confirming elevated cerebral CD163 shedding.

**Fig. 1.**
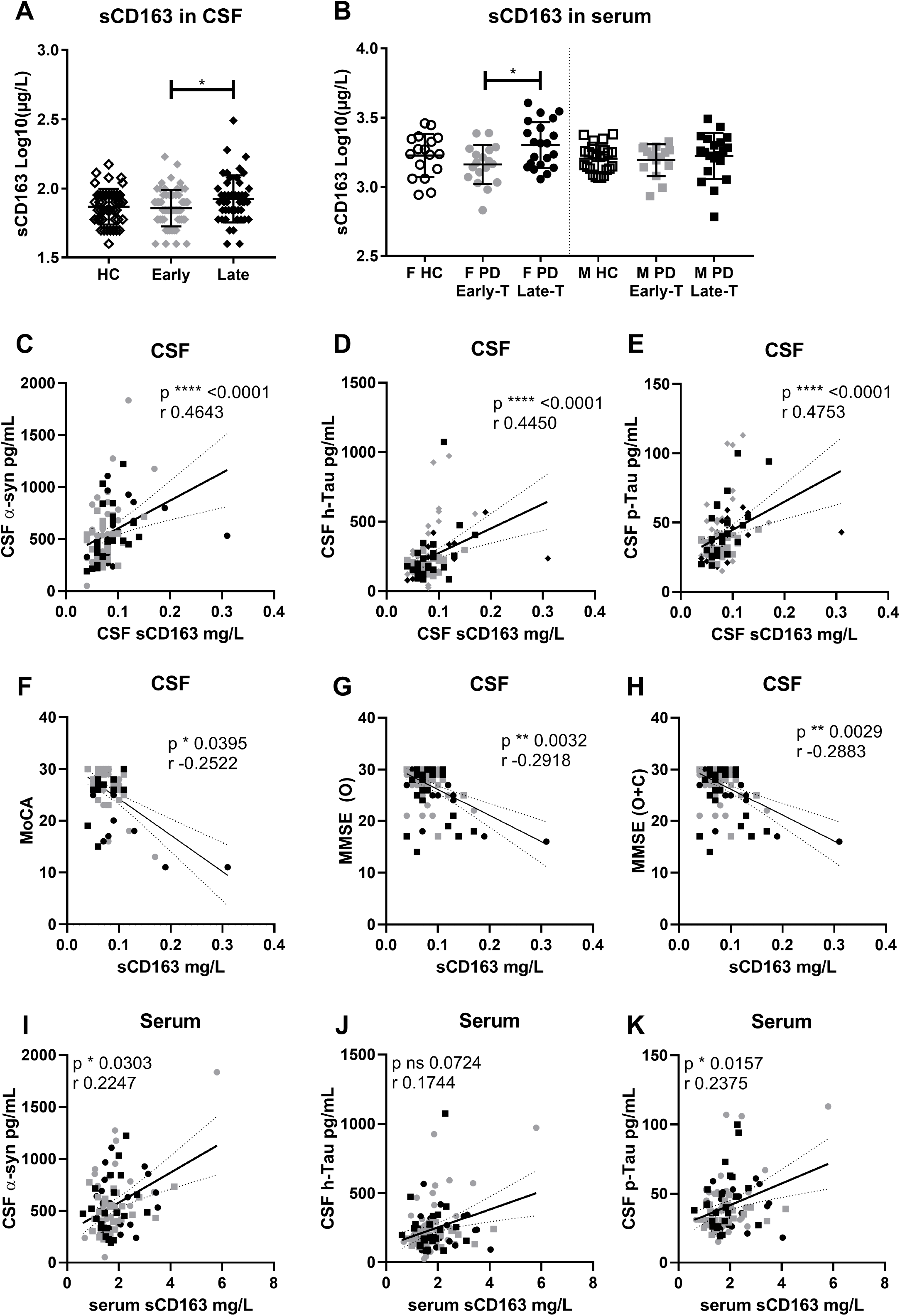
Identification of sCD163 as a biomarker in PD (Exp#1). Soluble (s)CD163 levels in CSF **A)** and serum **B)** from healthy controls (HC) or Parkinson’s disease (PD) patients at early (<5 years) or late (≥5 years since diagnosis) stage. Serum measurements were separated into female (F) and male (M) groups due to a priori identification of sex difference, and include only treated patients. Data are mean ±SD, and values were log-transformed to achieve normality. CSF-sCD163 for all PD patients correlated with well-characterized neurodegenerative CSF markers: **C)** alpha-synuclein (α-syn), **D)** total Tau (h-Tau), and **E)** phosphorylated Tau (p-Tau); as well as with clinical cognitive scores: **F)** the Montreal Cognitive Assessment (MoCA), **G)** the Mini-Mental State Exam (MMSE) original score (O), or **H)** original plus converted from MoCA (O+C). **I)** CSF α-syn, **K)** p-Tau and serum-sCD163 were correlated and showed a similar trend for CSF levels of **J)** h-Tau. P values for one-way ANOVA with Tukey’s multiple comparisons test values, respectively, Spearman two-tail p values (* <0.05, ** <0.01, ***<0.001, ****<0.0001), Spearman r, and best-fit slope with 95% confidence intervals are plotted.

Interestingly, for PD patients, CSF-sCD163 correlated positively with well-characterized neurodegenerative CSF markers (α-syn, h-Tau, and p-Tau), even after compensating for age and disease duration (**Fig.1C-E, Suppl.Table4**). sCD163 also correlated with h-Tau (Spearman r=0.477 p<0.01) and p-Tau (r=0.36 p<0.05) in HC. Since these biomarkers have been related to PD symptomatology, we examined putative correlations between sCD163 and patients’ clinical scores. Remarkably, CSF-sCD163 was negatively correlated with the cognitive scores MoCA and MMSE in the PD patients (**Fig.1F-H**). Serum-sCD163 also correlated positively with neuronal CSF markers (**Fig.1I-K**), even after compensation for covariance (**Suppl.Table4**). However, serum-sCD163 did not correlate with cognitive scores. Neither serum- nor CSF-sCD163 correlated with other clinical parameters (UPDRS-III or LEDD) after compensating for co-variables (not shown).

### Confirmation of sCD163 as a biomarker in PD (Exp#2)

We aimed to confirm these findings in a second independent measurement with samples from additional individuals in the same biobank (Exp#2) with a 2-year time gap between experiments (**Suppl.Fig. 1, Suppl.Table2**). Indeed, we corroborated an increase in CSF-sCD163 in late PD in Exp#2 (**Fig.2A**). Again, the estimated amount of sCD163 in CSF with serum origin was similar across groups (late PD 0.011±0.002, early PD 0.014±0.002, HCs 0.009±0.002), while the intrathecally produced sCD163 increased in late PD (0.109±0.047) vs. early PD (0.071±0.018, p=0.004) and vs. HCs (0.074±0.023 p=0.02). This suggests increased cerebral sCD163 production. We also confirmed the sex difference observed for serum-sCD163 with an increase in female patients only (**Fig.2B**).

**Fig. 2.**
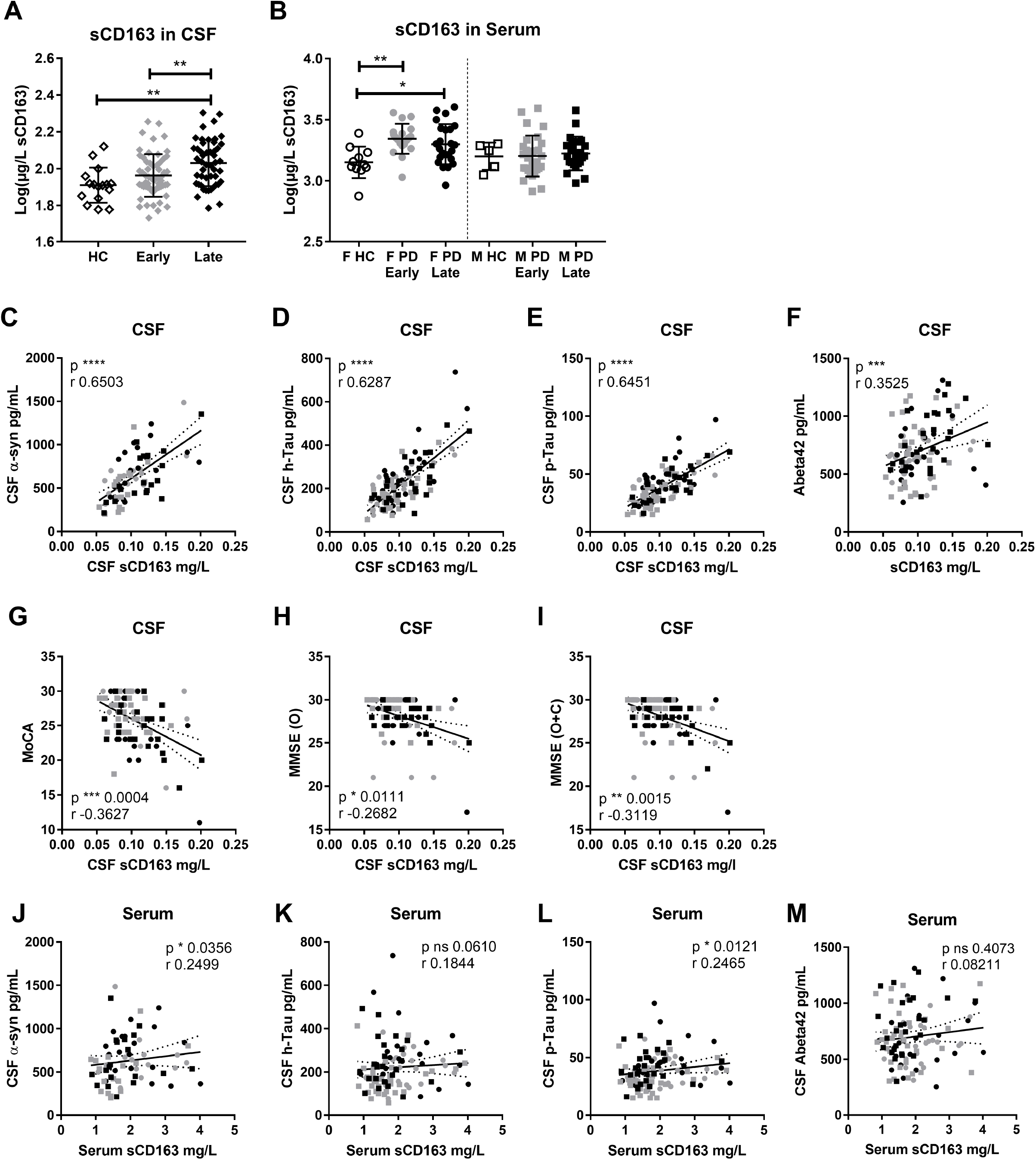
Confirmation of sCD163 as a biomarker in PD (Exp#2) Soluble (s)CD163 measurements in **A)** CSF and **B)** serum from healthy controls (HC) or Parkinson’s disease (PD) patients with early (<5 years) or late (≥5 years since diagnosis) status, with serum measurements being separated into female (F) and male (M) groups due to *a priori* identification of sex difference, and include only treated patients. Data are mean ±SD, and values were log-transformed to achieve normality. CSF-sCD163 for all PD patients correlated with well-characterized neurodegenerative/PD-CSF markers: **C)** alpha-synuclein (α-syn), **D)** total Tau (h-Tau), **E)** phosphorylated Tau (p-Tau), and **F)** Abeta42; as well as with clinical cognitive scores: **G)** the Montreal Cognitive Assessment (MoCA), **H)** the Mini-Mental State Exam (MMSE) original score (O), or **I)** original plus converted from MoCA (O+C). **J)** CSF α-syn, **L)** p-Tau and serum sCD163 were correlated and showed a similar trend for CSF levels of **K)** h-Tau. No correlation was found for serum sCD163 and **M)** Abeta42 in CSF, P values for ordinary or Kruskal-Wallis (non-parametric) one-way ANOVA with Tukey’s or Dunn’s multiple comparisons test values, respectively, Spearman two-tail p values (* <0.05, ** <0.01, ***<0.001, ****<0.0001), Spearman r, and best-fit slope with 95% confidence intervals are plotted. Further information on cognitive scoring correlations with respect to progression status and covariance can be found in **Suppl. Table 5**.

In agreement with observations from Exp#1, CSF-sCD163 correlated with the neuronal CSF markers (α-syn, h-Tau, and p-Tau) in Exp#2 (**Fig.2C-E**). CSF-sCD163 in HC also correlated with h-Tau (Spearman r=0.61 p<0.05) and p-Tau (r= 0.62 p<0.01). Moreover, in Exp#2, CSF-sCD163 was significantly correlated with levels of Abeta42 (**Fig.2F**). The association between serum-sCD163 and CSF biomarkers showed a similar pattern, with significant correlation with α-syn and p-Tau (**Fig.2J-M**).

Notably, this was also true for cognitive scores which decreased, while sCD163 increased in PD patients (**Fig.2G-I**). Age influenced cognition scores and sCD163; however, CSF-sCD163 levels remained significantly associated with cognitive decline after adjusting for age-at-visit alone, or in combination with age-at-onset or disease duration for the MMSE O+C scores (**Suppl.Table5**). Therefore, a 0.1mg/L increase in CSF-sCD163 corresponded to a decline of ∼3 MMSE O+C score. When adjusting for both age-at-visit and disease duration, the expected MMSE decline would be ∼1.8 (**Suppl.Table5**). This suggest that sCD163 is associated with cognitive decline, even when patient age and disease duration are considered.

### Other CSF and serum biomarkers (Exp#2)

To define the patients’ immune profile, we measured 40 immune-related serum and CSF biomarkers from Exp#2. In CSF, we observed that out of 16 reliable assays (**Suppl.Table3, Suppl.Fig2**), seven showed PD-related changes with a p<0.05: VCAM-1, ICAM-1, IL-8, SAA, PIGF, IL-15, and VEGF-D were increased in PD patients (**Fig.3A-K**). However, none remained significant after Bonferroni correction (α=0.0031).

**Fig. 3.**
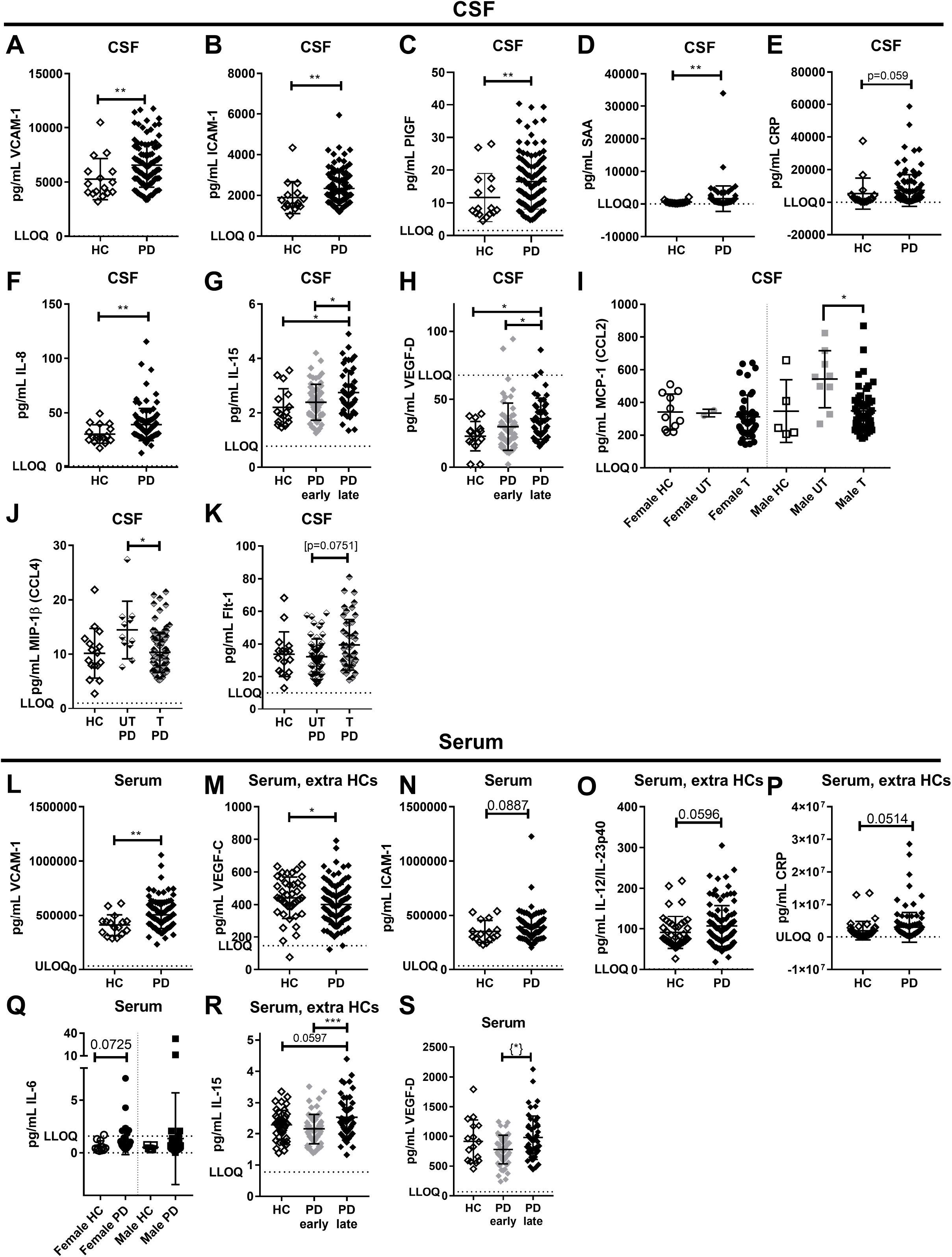
Screening of other CSF and serum biomarkers (Exp#2) Sixteen different CSF biomarkers and 27 serum biomarkers from the 40-plex mesoscale assay had values >LLOD (lower limit of detection) and were tested for differences **A-F&L-P)** between healthy controls (HC) and Parkinson’s disease (PD) patient groups; or **G-H&R-S)** between PD stages: early (<5 years) and late (≥5 years since diagnosis); or **I-K)** concerning treatment (T) status with or without (UT) L-dopa. **I)** MCP-1 (CCL2) and **Q)** IL6 were sex-separated due to a priori identification of sex differences. Twenty-six extra/additional HCs’ serum samples from Exp#1 were added for biomarkers unaffected by extra freezing/thaw cycle (**M, O, P, R**). Data are mean ±SD. P values are shown for unpaired t-test or Mann-Whitney test; for ordinary or (non-parametric) Kruskal-Wallis one-way ANOVA with Tukey’s or Dunn’s multiple comparisons test values, respectively, as appropriate: * <0.05, ** <0.01, ***<0.001, ****<0.0001. {*} one outlier from early PD was removed based on ROUT Q= 0.1%; if including the outlier: p=0.052. Lower and upper limit of quantification (LLOQ and ULOQ). Biomarkers with no differences (or no statistical trend) between groups are shown in **Suppl.Fig 2**.

In serum, only four markers (of 27 reliably assayed markers, **Suppl.Table3, Suppl.Fig2**) showed PD-related changes with p<0.05 (**Fig.3L-S**): VCAM-1 was increased in patients’ serum, and IL-15 and VEGF-D were both increased in late vs. early PD (**Fig.3L,R-S**), while VEGF-C was decreased in all patients (**Fig.3M**). Nevertheless, none remained statistically significant after Bonferroni correction (α=0.0018).

### Correlation between CSF and serum biomarkers with sCD163 (Exp#2)

To better describe the immune profile related to increased sCD163, we investigated any association with sCD163 and other immune molecules in CSF and serum (**Fig.4A-D**). CSF-sCD163 was positively correlated with several biomarkers in CSF, both when examining all PD patients and with early/late separation, even after Bonferroni correction. The strongest significant correlations for CSF-sCD163 observed at all disease stages were those with IL-15 (r 0.72) (**Fig.4G**), ICAM-1 (r 0.63), VCAM-1 (r 0.61), and Flt-1 (VEGFR1, r 0.51). Several chemokines were also strongly correlated with CSF-sCD163 in late PD: CXCL10 (r 0.52), CCL4 (r 0.51), and CCL2 (r 0.44) (**Fig.4A-B**).

**Fig. 4.**
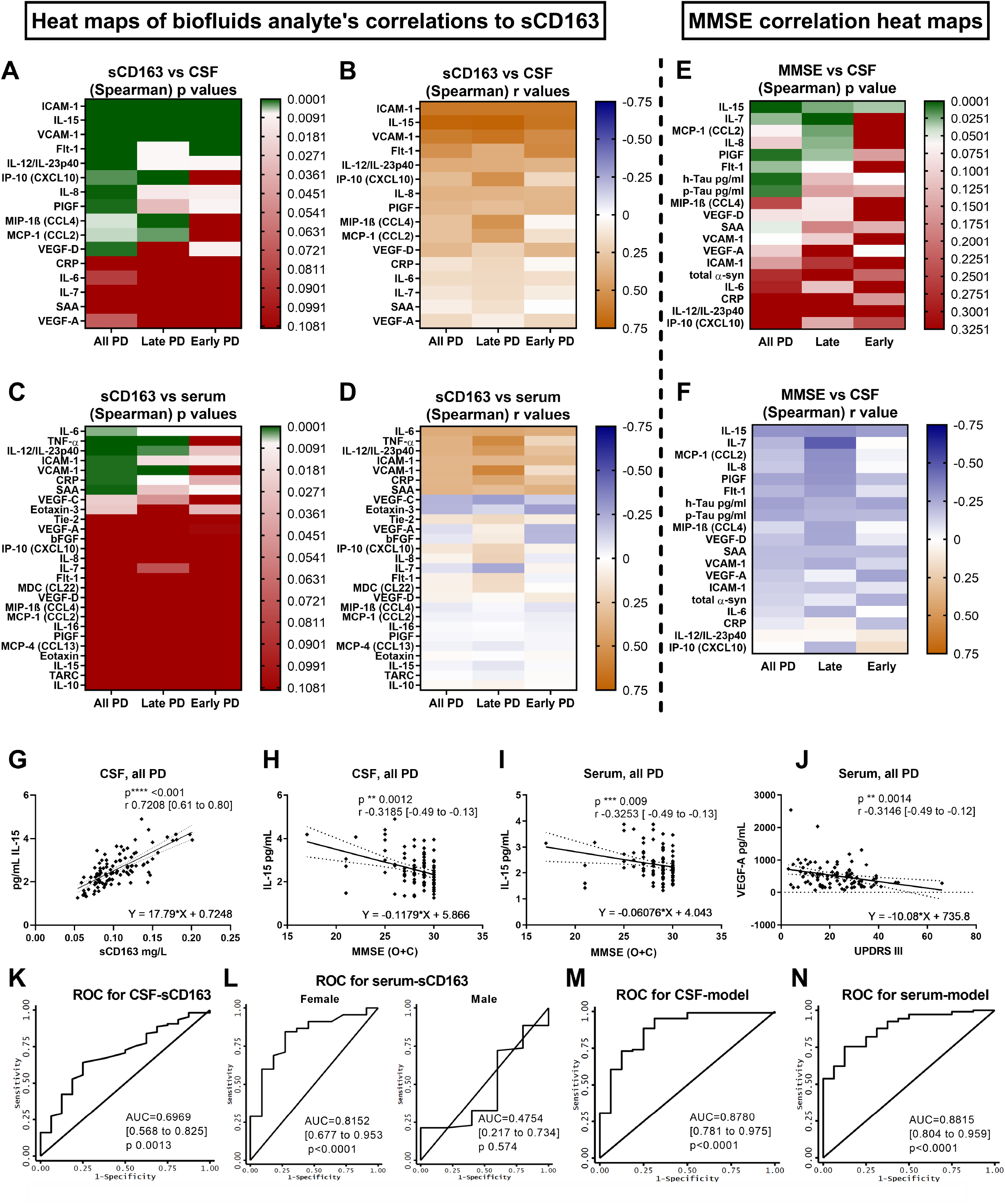
Correlations of sCD163 with other analytes, biomarker correlations with phenotypic scores, and ROC analysis of sCD163 and associated panels for PD diagnosis (Exp#2) **A-B)** CSF- and **C-D)** serum-sCD163 were compared with all 40-plex biomarkers with values above the lower limit of detection (LLOD) measured in CSF (n=16) and serum (n=27), respectively. **A&C)** Spearman correlation p value (the green color shows those with a p value below the Bonferroni-adjusted threshold [p<0.0031 for CSF and p<0.0018 for serum]) and **B&D)** Spearman r values are plotted as heat maps. Correlations are done for all Parkinson’s disease (PD) patients or separated as late (≥5 years since diagnosis) or early (<5 years since diagnosis) PD status. **E-F)** CSF biomarkers with values above LLOD are correlated with Mini-Mental State Exam (MMSE) (original scores + converted from Montreal Cognitive Assessment (MoCA) (O+C)) and plotted as heat maps for **E)** Spearman p and **F)** r values. Correlations are done for all patients with PD or separated as late (≥5 years) or early (<5 years since diagnosis) PD status. **C)** CSF IL-15 had the strongest correlation with MMSE O+C; plotted with the Spearman p and r [95% confidence interval (CI)] and the equation for the linear regression with a slope significantly different from 0. **D)** IL-15 was the only biomarker in serum with a significant correlation and linear regression with MMSE O+C. **E)** IL-15 in CSF had a strong correlation and linear regression with CSF-sCD163. **F)** Serum VEGF-A was the only biomarker with a significant correlation and linear regression with the Unified Parkinson’s Disease Rating Scale three (UPDRS III) score. Spearman two-tail p values (* <0.05, ** <0.01, ***<0.001, ****<0.0001). **K-N)** ROC curves for prediction of PD related to sCD163: **K)** CSF-sCD163 alone, **L)** serum-sCD163 alone when separated by sex, **M)** CSF M3 biomarker panel including sCD163, h-Tau, Abeta42, SAA, and VEGF-A x10, **N)** serum M3 biomarker panel including sCD163, VEGF-D/100, VCAM-1/10,000, Flt1/100, IL-15, IL-10, and MIP1β/100. Area under the ROC curve (AUC), confidence interval (CI). ROC curves for other biomarkers with p < 0.05 are shown in **Suppl.Fig. 3**.

In serum, the sCD163 correlations were weaker and fewer (**Fig.4C-D**): Serum-sCD163 was correlated with CRP (r 0.41), IL-6 (r 0.39), ICAM-1 (r 0.34), and SAA (r 0.35) in all patients, and with TNF-α (r 0.55), IL-12/IL-23p (r 0.47), and VCAM-1 (r 0.57), correlating at late PD. The strong sCD163/IL-15 correlation observed in CSF was absent in serum due to sex discrepancies in CD163 and not in IL-15.

### Phenotypic related biomarkers (Exp#2)

In CSF, besides sCD163, only IL-15 had a strong negative correlation with MMSE (O+C) and a significant linear regression (**Fig.4E-F,H**) as expected, since sCD163 and IL-15 in CSF were strongly correlated (**Fig.4G**). Interestingly, IL-15 was the only serum biomarker found to negatively correlate and have a significant linear regression with MMSE (O+C) (**Fig.4I**). Other CSF markers also showed a significant negative (but weaker) correlation with cognitive scores MMSE (O+C), although only in late PD: IL-7 (r −0.48), CCL2 (r −0.33), IL-8 (r −0.32), and PIGF (r −0.31) (**Fig.4E-F**). Only serum-VEGF-A showed a significant negative correlation and linear regression with UPDRS III scores (**Fig.4J**). Accordingly, serum-VEGF-A displayed UPDRS III prognostic potential in our mathematical analysis (see below, **Suppl.Fig. 5A**). No CSF biomarker correlated with UPDRS III (not shown).

### Mathematical modeling of sCD163 as phenotypic biomarker in PD (Exp#2)

In order to statistically evaluate the potential of sCD163 and the other assayed markers to predict PD diagnosis (PD vs. HC) and PD phenotypic scores (UPDRS-III, MoCA, MMSE O, and MMSE O+C), we used receiver-operating characteristic (ROC) analysis (area under the curve (AUC) estimates) and linear regression (regression slopes) (**Suppl.Fig. 3,5-6**). In CSF: sCD163, VCAM-1, ICAM-1, IL-15, IL-8, SAA, PIGF, and VEGF-C; and in serum: sCD163 (female), CRP (female), and VCAM-1 (all) showed statistical power to predict PD (**Fig.4K-L, Suppl.Fig3**). Principal component analysis (PCA) of CSF data confirmed sCD163, VCAM-, ICAM-1, IL-15, and IL-8 as markers associated with PD diagnosis (**Suppl.Fig4**). In CSF, the best individual markers to predict scores of all three cognitive scales used were sCD163, SAA, and IL-15; while in serum, they were SAA, CRP, and IL-8 (**Suppl.Fig. 5-6**).

Therefore, our analysis showed the prognostic power of serum- and CSF-sCD163 for PD diagnosis and cognitive scores (in CSF only). However, the specificity and sensitivity of sCD163 were not enough to be used alone. Therefore, we aimed to compute combined biomarker panels to address PD diagnosis and phenotypic scores using three variants of *stepwise forward modeling* with variable probability for biomarkers to enter the model (M), and with sCD163 being forced in M3 (**Suppl.Table6&7**). The best prediction model of serum and CSF biomarker panels were selected and plotted as PD prediction ROC plots (**Fig.4M-N)** or as goodness-of-fit plots for PD phenotypic scores (**Suppl.Fig. 7**). M3 including sCD163 together with other independently contributing biomarkers resulted in the best prediction of PD diagnosis, both in serum and CSF (both AUC 0.88). Likewise, M3 including sCD163 (in serum together with IL-15, CRP, CCL2 (MCP-1), PIGF, and IL-7; and in CSF together with p-Tau, IL-7, and IL-8) resulted in the best prediction of MoCA scores with the highest R-square values (serum 0.29, CSF 0.32) (**Suppl.Fig. 7B&F**). Therefore, sCD163 in serum and CSF holds promising potential as a significant contributor in a biomarker panel to address PD diagnosis, and cognitive scores, and deserves future investigation.

### Exploring *in vitro* interaction between CD163 and α-synuclein

Since sCD163 positively correlated with α-syn in CSF, we sought to further investigate any possible relation between them *in vitro*. No signs of direct co-interaction were observed using MST in any condition assayed (**Suppl.Fig. 7**). However, incubation of human primary MDMs with increasing concentrations of monomeric and fibrillar α-syn induced macrophage activation and dose-dependent sCD163 shedding (**Fig.5A**), supporting the inflammatory capacity of α-syn and its ability to induce CD163 cleavage, as seen for LPS (positive control).

**Fig. 5.**
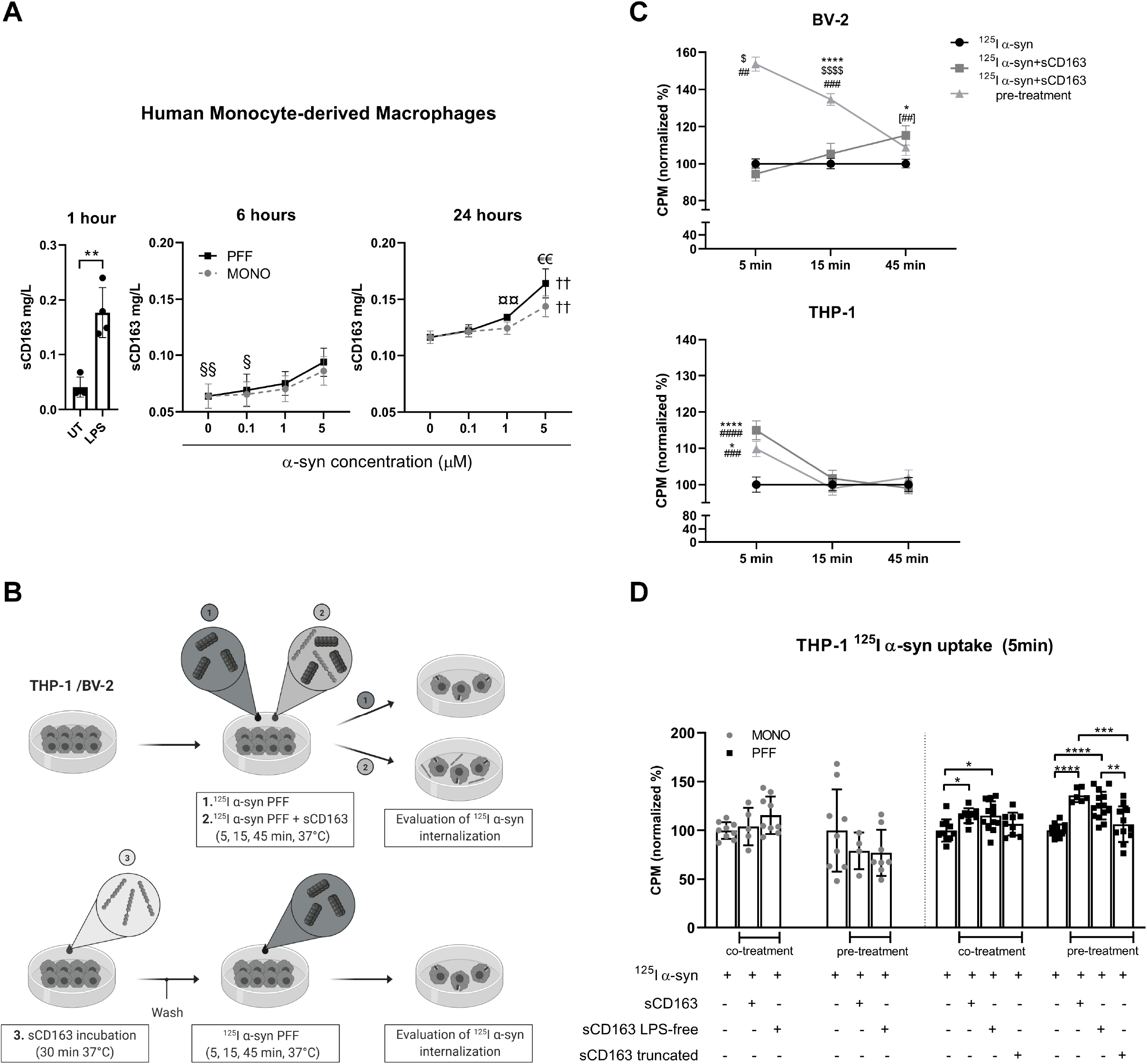
sCD163 increases α-syn uptake in myeloid cell lines; and α-syn induces sCD163 shedding in primary MDMs. The interaction between α-syn and sCD163 was studied *in vitro* **A)** sCD163 shedding in primary human monocyte-derived macrophages from four human donors after stimulation with LPS (positive control) or dose-dependent stimulation (0.1µM, 1µM, or 5µM) with monomeric (MONO) or pre-formed fibrils (PFF) α-syn for 1, 6, or 24 hours, respectively. **B)** Experimental design used in **C**): BV-2 and differentiated THP-1 cells were incubated at three different conditions: **1**. With iodine-radiolabeled α-syn (^125^I α-syn) PFF (control); **2**. Co-incubated with ^125^I α-syn and sCD163 (5μg/mL); and **3**. Pre-treated with sCD163 (5μg/mL) 30 min prior ^125^I α-syn addition. Cells were incubated at 37°C (5, 15, and 45 min) after α-syn addition and collected for evaluation of α-syn internalization. Radioactive counts per minute (CPM) on differentiated THP-1 and BV-2 cells were normalized to α-syn average counts (treatment 1) and shown as % of control internalization/binding. Endotoxin levels in recombinant sCD163 were measured and corresponded to 0.030EU/mL (according to concentration used). **D)** The sCD163-mediated uptake of MONO and PFF α-syn by THP-1 cells was evaluated after 5 min of incubation of full-length (domain 1-9) “LPS-free” sCD163 (LPS-removal treatment on the sCD163 protein sample resulting in 0.00017EU/mL endotoxin, according to concentration used) or a truncated version (domain 1-5) of sCD163, where LPS was also removed. Statistics: Paired t test; **A-D)** two-way ANOVA followed by post-hoc Tukey’s multiple comparison test when appropriate. P values are given in intervals according to the number of symbols, e.g. *<0.05, **<0.01, ***<0.001, ****<0.0001. Symbol explanation: **A)** § different from 5µM in PFF; † different from all other in the same group; € PFF different from MONO in the same dose; ¤ different from 0µM in PFF. **A-D)** * different from control (**A)** UT or **C-D)** α-syn alone) at the same time point. **C)** $ different from all other treatments at the same time-point; # different from adjacent time point in the same treatment; [##] co-treatment different from 5 min. Data are shown as mean ± SEM. Illustration was made using Biorender.

We then evaluated the effect of sCD163 on α-syn uptake using human monocytic THP-1 cells differentiated into macrophages and mouse microglia BV-2 cells to obtain information from the two myeloid cells involved in PD. Cells were incubated with radiolabeled α-syn fibrils alone or with recombinant sCD163 as co- or pre-treatment (30 min) (**Fig.5B**). After 5 min, THP-1 cells increased α-syn internalization when co- or pre-treated with sCD163 compared with α-syn alone. In BV-2 cells, the uptake of α-syn increased after 5 and 15 min of pre-treatment with sCD163, while co-incubation increased α-syn uptake only after 45 min (**Fig.5C**). This increased α-syn fibril uptake in THP-1-derived macrophages was specifically associated with full-length sCD163 since LPS removal did not affect uptake, whereas truncation of the sCD163 protein did (**Fig.5D**).

## DISCUSSION

Genetic data suggest that the PD inflammatory component is driven by the myeloid immune compartment, which includes microglia and monocytes/macrophage^28^. Numerous studies show alterations on immune biomarkers in CSF and serum associated with symptoms in PD patients^11^. However, these biomarkers are not cell type specific and therefore provide no information on cellular relevance in the disease mechanism. Here, we show that CSF-sCD163, a monocyte/macrophage-specific biomarker^14^, increases in late vs. early PD, hence suggesting increasing monocytic activation with disease duration. sCD163 was associated with well-accepted PD biomarkers and inversely correlated with cognitive scores, supporting a role for monocytes in neurodegeneration. Serum-sCD163 was also increased, although only in female patients, suggesting a sex-distinctive monocytic response. This increase in sCD163 levels is indicative of monocytic activation occurring in parallel with variations in cytokines, chemokines, and angiogenic factors. These changes were different in CSF vs. serum, supporting a distinctive immune profile in the brain and periphery. Step-forward regression models and ROC analysis confirmed sCD163 as a predictor of PD and cognitive scores. Additionally, mathematical models confirmed the prognostic potential of sCD163 in combination with other biomarkers in serum and CSF. Interestingly, our *in vitro* studies suggest that α-syn itself can induce macrophage activation and sCD163 shedding, while sCD163 might enhance α-syn uptake by myeloid cells and participate in the clearance of α-syn. In conclusion, our data suggest that sCD163 is a promising biomarker associated with inflammation in PD, thereby supporting a role for α-syn in monocyte activation and sCD163 shedding that could have direct consequences for α-syn processing.

Our analysis of the two experiments in the PD cohort showed increased sCD163 in biofluids from PD patients. CD163 is cleaved by ADAM17^22^, which also cleaves VCAM-1 and ICAM-1^29, 30^. ADAM17 activity is enhanced by inflammatory signals, such as TLR2/4 activation; therefore, sCD163 levels are related to monocyte activation and associated with severity of diseases with an inflammatory component^14^. Indeed, sCD163 is widely used as an inflammatory marker for disease prognosis in multiple disorders including diabetes, asthma, and atherosclerosis^31^. Two diseases genetically related to PD also show increased sCD163: Crohn’s syndrome^32^ and Gaucher disease^33^, confirming a shared common myeloid disease mechanism. The inflammatory signals responsible for CD163 shedding in PD are yet unknown, but α-syn may be a contributor since it can activate TLR2/4^2^, which are upregulated in PD patients’ monocytes^4, 34^. Accordingly, we demonstrated sCD163 shedding in human primary MDMs incubated with fibrillar α-syn, confirming the inflammatory ability of α-syn in macrophages^35^ and its relation to the increase of sCD163 in PD.

Our data from this cross-section cohort showed increased sCD163 with time from onset, supporting increasing involvement of CD163^+^ monocytes in the different PD stages. Higher CSF-sCD163 levels were not a simple leakage from serum, since the increase was not found in male serum. Furthermore, the estimated amount of sCD163 produced intrathecally was, indeed, elevated in PD. The sCD163 found in CSF can be produced by CD163-expressing meningeal, choroid plexus and perivascular macrophages, as well as by infiltrating macrophages^17-19^. CD163 is a myeloid marker whose expression is lost during microglia development and is therefore absent in adult microglia, as recently shown in single-cell RNA studies^15-17^. In studies conducted by others and ourselves, an increase in CD163^+^ cells has been observed in brain parenchyma in rodent PD models^9, 10^ and *post mortem* brains of patients with Alzheimer and PD^36^. Thus, sCD163 could be partially produced by infiltrating CD163^+^ monocytes/macrophages and their subsequent local activation. Accordingly, CSF-sCD163 was correlated with CCL2, CCL4, and CXCL10, chemokines involved in monocyte recruitment and previously correlated with PD symptoms^37-40^. More importantly, sCD163 was also strongly correlated with regulators of angiogenesis, BBB extravasation, and/or monocyte brain-infiltration or recruitment^41-45^, which have been reported to be increased in PD ^46-48^-namely: ICAM-1, VCAM-1, and VEGF-D in both serum and CSF; and IL-8, IL-15, and PIGF only in CSF.

sCD163 is a biomarker widely used to measure an inflammatory component. Accordingly, in PD serum, sCD163 was associated with an increase in the acute phase proteins SAA and CRP, which were previously associated with motor impairment and PD dementia ^37, 49-51^. Moreover, serum-sCD163 also correlated with the pro-inflammatory cytokines TNF-α and IL-6, as well as IL-12/IL-23p40 and IL-15, both elevated in PD serum. IL-12/IL-23p40 is produced by dendritic cells (also CD163^+^) upon TLR activation, whereas IL-15 induces dendritic cell differentiation; thus, both are involved in T cell modulation^52,53^. IL-12/IL-23p40 was previously found to be increased and important in PD and Crohn’s disease^54, 55^. Taken together, patients’ CSF-sCD163 correlated with immune molecules associated with angiogenesis, cell-infiltration, and T cell activation, relating monocyte activation to leucocytes recruitment to sites of inflammation in the brain, whereas serum-sCD163 correlations indicated monocyte activation in relation to pro-inflammatory events. Overall, the immune profile in PD suggested occurrence of an inflammatory events leading towards a permissive environment for recruitment of adaptive and innate immune cells to the brain.

Surprisingly, IL-15 was the only biomarker elevated in PD serum and CSF correlating with cognitive scores in both biofluids, thus revealing that IL-15 may be another putative biomarker. However, despite the relevance of IL-15 in BBB permeability, brain immunomodulation, and its connection to mood and memory^56^, our ROC analysis for IL-15’s ability to predict PD diagnosis had a lower AUC with higher variation than sCD163. Moreover, in contrast to CD163, IL-15 is produced by a variety of cells including neurons, and is therefore not monocyte-lineage specific like sCD163^56^. Therefore, our data suggest a relevance of IL-15 in PD, but it does not provide information on specific cell types involvement in PD and cognition, and do not reflect the sex difference in the disease, which might be both a disadvantage and a gain.

Serum-sCD163 levels were only significantly changed in female PD patients. To our knowledge, only urate has been proposed as a PD biomarker with sex relevance^57^. Sex differences in the immune system are long acknowledged^58^. PD has a sex bias, and immune-related sex differences have been shown in PD^54^. Sex-specific changes in serum-sCD163 could be related to female-male differences in PD incidence^59^ and symptomatic presentation^60^. Accordingly, we previously showed sex-dependent variation in the response of PD blood monocytes *in vitro*^3^. Therefore, the observed differences could be a result of sex-specific immune divergences with consequences for the risk and presentation of PD.

CSF-sCD163 was strongly correlated with the disease biomarkers α-syn, h-Tau, and p-Tau, which have been associated with PD diagnosis, clinical symptoms, and cognition^61, 62^. Our data suggest that in PD, sCD163 increases with α-syn. This correlation could be directly related to the ability of α-syn to induce monocytic activation and sCD163 production, since α-syn has been reported to longitudinally increase with time in PD^63^. Similarly, h-Tau and p-Tau also increase, probably reflecting progressive neurodegeneration.^63^ We found that higher sCD163 levels were associated with lower cognitive scores, supporting a driving role for monocytes in the cognitive component of PD. This agrees with imaging studies showing that cognitive scores in PD dementia correlated with increased immune activation and reduced cortical glucose metabolism^64^. Furthermore, changes in monocytes seem more relevant in those PD patients with higher risk of developing cognitive problems^4^. Altogether, our data support a relation between monocyte activation and the PD cognitive component.

sCD163 function is unclear, but it was shown to potentiate IL-10 expression in allergen-activated PBMCs^65^ and to inhibit T cell activation/proliferation^66^, hence promoting inflammatory resolution. CD163 facilitates hemoglobin-haptoglobin complex^67^ and ADAMTS13^68^ internalization. Our analysis showed no direct binding between CD163 and α-syn, regardless of calcium concentration, suggesting that α-syn is not a direct ligand for CD163 (possible co-receptor interactions cannot be excluded). However, our *in vitro* study suggests that α-syn can induce monocyte activation and sCD163 shedding, which in turns increases the capacity of monocytes and microglia to uptake extracellular α-syn. *In vitro*, T cell-internalized sCD163 binds myosin^69^, a molecule involved in phagocytosis^70^. Therefore, sCD163 might induce intracellular pathways that would influence α-syn phagocytosis and thus resolve inflammation. Indeed, sCD163 required time to exert its effect on BV-2 cells, supporting induction of certain intracellular cascades. Otherwise, this delay could be due to species disparities. Further investigation is required to fully understand the sCD163 effect on myeloid response to α-syn during PD.

The findings of the present study may be somewhat limited by the cross-sectional study design, low numbers of HCs, and potential cofounding inflammatory diseases (with anti-inflammatory medication) among study participants. The increase in sCD163 in late PD compared to early PD and HCs, as well as the negative correlation with cognitive scores suggest a relation between sCD163 and PD stages. However, we cannot exclude a possible relation with sCD163 and cognitive decline during ageing in other synucleinopathies and neurodegenerative diseases. Nevertheless, our mathematical modelling proposes a prognostic power for biomarker panels including sCD163 as one of the analytes. However, this has to be examined in a prospective longitudinal study to truly determine the potential of sCD163 as a PD and cognition biomarker. In conclusion, we show a PD-related increase in sCD163 in serum (in females) and CSF, supporting a role for monocytes in the PD immune response. The increase in sCD163 paralleled those of accepted neuronal biomarkers, therefore relating monocytic and neuronal events. Additionally, higher sCD163 levels were associated with lower cognitive scores, indicative of an association between the monocytes’ immune response and cognitive decline. The CSF-sCD163 increase was associated with immune biomarkers, suggestive of a permissive BBB and activation of adaptive immune cells. Moreover, our *in vitro* studies suggest that α-syn could activate myeloid cells and induce sCD163 release, which enhances the myeloid cells’ capacity to uptake α-syn, indicative of a relevant role for disease progression.

## Data Availability

Qualified investigators can request raw data through corresponding author.

## ACKNOWLEDGEMENTS

We acknowledge the invaluable technical help provided by Gitte Ulbjerg Toft and Gitte Fynbo Biller (Department of Biomedicine, Aarhus University) and by Helle Hauser Ryom and Christina Strande Søndergaard (Department of Clinical Biochemistry, Aarhus University Hospital). We are grateful to Prof Dr. Mart Saarma (Helsinki University) for his scientific support and Prof Poul Henning Jensen (Aarhus University) for kindly providing the human α-syn protein. Samples were obtained from the Neuro-Biobank of the University of Tuebingen, Germany (https://www.hih-tuebingen.de/en/about-us/core-facilities/biobank/), which is supported by the local University, the Hertie Institute and the DZNE.

## AUTHOR CONTRIBUTIONS

Developed the concept and designed the study: SKN, SAF, MRR. Performed experiments: SKN, SAF, DH, AP, MCN, JHG, AE. Analysed the data: SKN, SAF, KS, AP, MRR. Drafted the manuscript: SKN, MRR. Coordinated sample selection and collected biobank archive info: CS. Wrote jointly the final version of the manuscript: SAF, SKN, MRR. All authors critically revised the manuscript and approved the final version.

