## Supplementary info for "“Soluble CD163 changes indicate monocyte association with cognitive deficits in Parkinson’s disease”"

**Participants and clinical analysis**

Parkinson's disease (PD) patients were recruited from the outpatient clinic and/or ward for Parkinson's disease at the University Hospital of Tuebingen. Spouses of patients and volunteers recruited by newspaper advertisements were assessed to have no neurodegenerative disease and served as controls. All participants were examined by a neurologist specialized in movement disorders. Diagnosis of PD was made according to the UK Brain Bank Society Criteria [1]. None of the included patients showed "red flags" indicating an atypical parkinsonism over time. PD patients were assessed in the dopaminergic ON state. We assessed severity of motor symptoms using part III of the MDS Unified Parkinson's disease Rating Scale (MDS-UPDRS III) [2]. Disease stage was categorized by the modified Hoehn and Yahr Scale (H&Y) [3]. Cognitive function was tested using the Montreal Cognitive Assessment (MoCA) [4] and/or the Mini Mental Status Examination (MMSE) [5]. Since the MoCA was available only from 2009 on, all previously obtained MMSE scores were converted into MoCA equivalent scores according to a published algorithm [6]. Cognitive impairment was defined *according to the DSM-IV (2003-2013) or DSM-V (2013-2019) criteria and/or MoCA ≤25 as reported* by Hoops et al. (point of maximum combined sensitivity and specificity) [4]. Mood disturbances were assessed using Beck's Depression Inventory II (BDI II) [7]. The study was approved by the local ethics committee (480/2015BO2), with all participants providing informed consent.

Cofounding inflammatory diseases were not informed and thus not corrected for, nor the use of anti-inflammatory medication. However, a list of immune related drugs was extracted from the biobank archives and using Fishers exact test, we did not observe any significant difference in the number of persons taking anti-inflammatory drugs between the PD and HC group in both Exp#1 and Exp#2. Specifically, anti-inflammatory medications used were as follows: Exp#1: aspirin (2 male-HC, 1 female-PD, and 5 male PD) ibuprofen (2 female-PD (1 using also dexamethasone) and 1 male-PD) or diclofenac (1 female-PD). For Exp#2: for aspirin (3 female-PD and 8 male-PD), ibuprofen (2 female-PD and 1 male-PD), diclofenac (1 male-PD) and budesonide (1 male-PD, who was using also Insulin). Overall, in Exp#1: 2 HC vs. 10 PD; and in Exp#2: 0 HC vs. 16 PD were using some anti-inflammatory medication. Anti-inflammatory specific for digestive tube related diseases: Exp#1: loperamide (1 female-PD) and mesalazine (1 male-PD, samples from this male are also included in Exp#2).

**Collection of CSF and serum Samples**

Spinal tap for CSF collection along with serum collection were performed between 9.00 AM and 1.00 PM. Samples were directly taken from the bedside and centrifuged within 60 min after collection and frozen at -80°C within 90 min after collection. Samples with abnormal routine CSF diagnostics (white blood cell count >6 cells/µl, Immunoglobulin Subtype G (IgG) index > 0.7) were excluded.

**Biomarkers measurements**

sCD163 was measured in serum and CSF using an in-house ELISA[8, 9]. The CSF concentration of sCD163 derived from the circulation was calculated based on the CSF/serum albumin quotient: serum sCD163 x (CSF-albumin/serum-albumin concentration). The intrathecally produced sCD163 was calculated by subtracting the above value from the absolute CSF-sCD163 concentration[10].

ELISA was used to measure biomarkers in CSF: Total human Tau (h-Tau) and phosphorylated Threonine 181-Tau (p-Tau) (Innotest; Innogenetics, Ghent, Belgium); and $Abeta_{1-42}$ (Abeta42) (Innotest; Fujirebio Germany GmbH, Hannover, Germany). Total α-syn values were analyzed at Analytic Jena, (Mono-Kit, Analytic Jena Roboscreen GmbH, Leipzig, Germany). All measurements were performed by board-certified laboratory technicians who were blinded to clinical data.

**40-plex mesoscale**

MSD MULTI-SPOT Assay System (Mesoscale) was used to measure 40 different soluble molecules in serum and CSF on Exp#2 (and additional serum controls from Exp#1, see **Suppl. Fig. 1, Suppl. Table**

**2&3**), using Human Biomarker 40-plex kit containing five different V-PLEX kits following manufacture's best practices instructions: Pro-inflammatory Panel 1 (human) [IFN-γ, IL-1β, IL-2, IL-4, IL-6, IL-8. IL-10, IL-12p70, IL-13]; Vascular Injury Panel 2 (human) [SAA, CRP, VCAM-1, ICAM-1]; Chemokine Panel 1 (human) [Eotaxin, MIP-1β, Eotaxin-3, TARC, IP-10, MIP-1α, IL-8, MCP-1, MDC, MCP-4]; Cytokine Panel 1 (human) [GM-CSF, IL-1α, IL-5, IL-7, IL-12/23p40, IL-15, IL-16, IL-17A, TNF-β, VEGF-A]; Angiogenesis Panel 1 (human) [VEGF-A, VEGF-C, VEGF-D, Tie-2, Flt-1, PIGF, bFGF]. Due to different optimal concentration quantification ranges for the double measurement of IL-8 and VEGF-A in the two different kits, they were only calculated based on Pro-inflammatory Panel 1 and Angiogenesis Panel 1, respectively. Calibrators and a plate-plate control (mixed human serum frozen in several aliquots) were run in duplicates and used for %CV variance validation within and among plates (cut off >25%). Serum and CSF samples were separated on different plates due to different (pre-) dilutions. Patients' samples were run as singlets, with the different patient and control groups evenly distributed on two plates per panel.

All samples were used after a single freezing cycle, except for the additional healthy serum controls from Exp#1 (**Suppl.Fig.1/Suppl.Table2**), which had been thawed once before, and were only included in assays where we had first validated that the additional freeze/thawing-cycle did not affect the measurements (**Suppl.Table3**). Briefly: Angiogenesis panel 1 was blocked for 1 hour with shaking at room temperature (RT), samples were thawed on ice and centrifuged at 2000xg for 3 min to avoid debris/precipitations. Calibrators were mixed in appropriate diluents in Eppendorf tubes. Samples and calibrators were aliquot or diluted in polypropylene 96 well plates (Vascular injury panel 2: serum 20x+50x and CSF 5x). V-PLEX plates were washed 3 times in wash buffer (PBS + 0.05% Tween20); calibrators, diluents, and samples were transferred to plates for a final volume of 50µL 2x (Pro-inflammatory, Angiogenesis and Cytokine Panel 1) or 4x (Chemokine panel 1) dilution (except for Vascular injury panel 2: 25µL, and no further dilution). V-PLEX plates were incubated at RT at 2 hours while shaking followed by 3 washes, and incubation with 25µL detection antibody (AB) mix for 1 hour (Vascular injury panel) or 2 hours (all other panels) while shaking at RT. After 3 washes, 150µL 2x Read Buffer T was added and incubated for 3 min (except Chemokine panel 1: 10min) and read using electrochemiluminescence on MESO QuickPlex SQ120 (MSD) with MSD Discovery Workbench software. For each plate and panel, the upper- and lower limit of quantification (ULOQ & LLOQ) or detection (ULOD & LLOD) were noted. No measurements were >ULOD, only assays with <3 measurements below LLOD were considered reliable and further analyzed (**Suppl. Table3**). ULOQ or LLOQ are shown on relevant graphs.

**Production of full-length sCD163 and endotoxin measurement**

cDNA encoding hCD163 short tail variant (Uniprot-KB Q86VB7-3) with an insert encoding the amino acid sequence HHHHHHDDDDK between Gly46 and Thr47 was cloned into the pcDNA3.4 vector (Invitrogen Geneart, Thermo Fisher Scientific, Regensburg, Germany) and transiently expressed in the ExpiCHO$^{TM}$ expression system (Gibco, Thermo Fisher Scientific) according to manufacturer's instructions. 30mL supernatant was harvested and shedded CD163 was purified on a 5ml Ni-NTA Superflow column (Qiagen) with 1ml/min flow. The column was washed with buffer A for 6 CV and eluted with a gradient from 0-100% over 15CV (buffer A: 50mM $NaH_2PO_4$, 300mM NaCl pH8.0; buffer B: 50mM $NaH_2PO_4$, 300mM NaCl, 250mM Imidazol pH8.0). Purified protein was buffer exchanged into a 150mM NaCl and 10mM Hepes pH 7.5 buffer.

Endotoxin levels in purified protein preparations were measured using the HEK-Blue cell based LPS detection method (Invivogen). In short, HEK-Blue-4 cells expressing human TLR4 and a NF-kB-inducible secreted embryonic alkaline phosphatase reporter (SEAP) were seeded in 96-well plates together with 1:10 to 1:2560 dilutions of purified protein or an endotoxin standard curve (1-0.004EU/ml) and incubated at 4°C 5% $CO_2$. After 24 hours, supernatant was harvested and level of endotoxin was determined by analyzing SEAP activity in the supernatant using 1mM 4-Methyl umbelliferone (Sigma Aldrich) as substrate. Then endotoxin level in the sCD163 protein solution was measured and the concentration used in the assay corresponds to 0.030EU/mL.

**Human CD163 SRCR 1-5 cell lines used for production of truncated version of CD163**

Stable transfected CHO cells expressing SRCR domain 1–5 of human CD163 were established by transfecting a CHO K1 cell line with a pEF4/V5-His transfection vector (Invitrogen, Taastrup, Denmark) containing cDNA encoding amino acids 1-578 of human CD163 in-frame with the V5-His6. In details, the transfection vector was generated by amplifying cDNA encoding amino acids 1-578 of human CD163 by standard PCR using full-length human CD163 cDNA as template and the following primers:

Fwd 5'-GAGGGTACCATGAGCAAACTCAGAATG

Rev 5'-CTCTTCGAATGAGCAGACTACTCCAAC.

Amplified cDNA was subcloned into pEF4/V5-His using KpnI and BstBI restriction enzymes and T4 ligase (Thermo Fisher). Cells were grown in HyClone SFM CCM5 with hygromycin (100μg/ml) and PEN/Strep using 300μg/mL Zeocin (Invitrogen) for clonal selection. Two liters of media supernatant was collected and Tris-HCl buffer pH 8.0 was added to a final concentration of 10mM. The supernatant was passed over 20mL NiNTA-agarose (Quiagen), washed with 300mM NaCl in 50mM Tris-HCl pH 8.0 and gradient eluted in that buffer with imidazole from 0 to 400mM. The fractions containing the CD163 domain 1-5 were buffer exchanged into 10mM NaCl in 25mM Tris-HCl pH 8.0, loaded on a 20ml Q-sepharose column, and gradient eluted from 10 to 500mM NaCL in 25 mM Tris-HCl pH 8.0. The fractions containing the CD163 domain 1-5 were pooled and buffer exchanged into PBS.

**LPS removal from sCD163 protein solution**

LPS was removed from purified truncated sCD163 and full-length sCD163 when noted as "LPS-free": Triton-X114 essentially was used as previously described [11]. In short, Triton-X114 (Sigma-Aldrich) was added to the sCD163 solution to a final concentration of 2%, incubated at 4°C for 30 min while stirring, followed by incubation at 37°C for 10 min while stirring, and subsequently centrifuged at 20,000g at 37°C for 20 min. The water phase (supernatant) containing the purified sCD163 protein was collected and gel-filtrated to ensure removal of residual Triton-X114. The remaining level of endotoxin in the "LPS-free sCD163" full-length protein solution was measured (see above) and the calculated concentration used in the assay was 0.00017EU/mL.

**Lactoperoxidase-catalyzed iodination of α-synuclein fibrils**

Human α-syn fibrils (a kind gift from Prof. Poul H. Jensen, Aarhus University) were produced as previously described [12], diluted to 0.1mg/ml concentration in PBS and sonicated in a bath sonicator (Bioruptor, NGS, Diagenode) using high power and 10 cycles with 30 sec on and 30 sec off. For iodination, 5μg of sonicated human α-syn fibrils in 50μl PBS were mixed with 50μl of 250mM phosphate buffer, pH 7.5; then, 1mCi of $^{125}$I Na (PerkinElmer), 10μl of lactoperoxidase solution (50μg/ml, Merck, Germany), and 10μl of 0.03% $H_2O_2$ in PBS (diluted from 30% solution, Merck, Germany) were added. The mixture was incubated in 37°C water bath for 20 min, and the reaction was stopped by adding 200μl of 0.1M phosphate buffer, pH 7.5, containing 0.1M NaI and 0.42M NaCl. Next, 25μl of 2.5% BSA in 0.1M phosphate buffer, pH 7.5, was added to the sample and loaded onto Sephadex G-25 column (PD10, GE Healthcare, UK) to separate free iodine from the labeled fibrils. The elute was collected into 500μl fractions, and 1μl of each sample was counted at the Wallac Gamma Counter (Wallac/LKB) to detect the fraction that contained the highest gamma-counts i.e. the labeled protein. The column had been equilibrated by eluting with 30-40ml of 0.1M phosphate buffer, pH 7.5, containing 1% BSA. The labeled $^{125}$I- α-syn fibrils were used fresh, within the following 6 hours, to avoid aggregation into larger species.

**Analysis of α-synuclein uptake and binding**

THP-1 and BV-2 cells were grown in 75cm$^2$ flasks in culture media consisting of 2mM L-Glutamine, 10% FBS in RPMI 1640. For the THP-1 experiment, 80.000 cells per well were plated onto 24-well plates and treated with 100nM PMA (Phorbol myristate acetate, InvivoGen, France) for 72 hours for macrophage-differentiation and attachment. For the BV-2 experiment, 70,000 cells per well were plated onto 24-well plates and cultured for 24 hours before α-syn treatment.

The plates with differentiated THP-1 macrophage-like cells or BV-2 murine microglia cells were placed on ice for 15 min before adding $^{125}$I-α-syn fibrils (50 ng/ml) onto the cells, either with PBS as vehicle control, sCD163 (5µg/ml), or added after 30 min of pre-treatment with sCD163 (5µg/ml) at 37°C. To evaluate α-syn internalization, the plates were placed into a 37°C water bath to allow uptake of the fibrils for 5, 15, or 45 min, after which the plates were returned on ice. The cells were washed with 500µl ice-cold PBS twice for 5 min and next, to remove the bound preformed fibrils from the cell surface, they were washed with 500µl acid wash (0.2M acetic acid, 0.5M NaCl, pH 2.8) for 5 min. Afterwards, the cells were lysed by adding 500µl 1M NaOH to each well and collected into appropriate counting vials. Radioactive counts per minute (CPM) were counted in the Wizard 3" 1480 Automatic Gamma counter (Wallac/LKB). For assessment of surface binding, the plates were kept on ice for 1 hour, washed with 500µl ice-cold PBS twice for 5 min, lysed, and collected with 500µl of 1M NaOH into vials for counting. Values were normalized to α-syn average counts. Results are given in percentage of internalization/surface binding.

**Microscale Thermophoresis for assessment of sCD163-α-synuclein interaction**

Evaluation of protein binding through the Microscale Thermophoresis (MST) technique was performed by labeling human His-Tag sCD163 (target protein) with RED-tris-NTA 647 2$^{nd}$ generation fluorescent dye (excitation:650nm; emission: 670nm) using the Monolith His-Tag labeling Kit RED-tris-NTA 2$^{nd}$ generation (NanoTemper Technologies, Cat Nr:L018). Fluorescently labeled sCD163 was used at a concentration of 60nM. For binding evaluation, a 16x dilution series (1:1) was performed on 20µM human α-syn (ligand) reaching 0.61nM concentration. The MST buffer used contained 10mM HEPES, 150mM NaCl, 0.05% Tween, 0.1% BSA, and three different concentrations of $CaCl_2$ for testing calcium-dependent binding. Fluorescent target and ligand were mixed (1:1) and samples were loaded in Monolith NT.115 Premium Capillaries (NanoTemper Technologies, Cat Nr: K025) and measured in the Monolith NT.115 instrument (100% LED/excitation power and medium MST power). The experiment was repeated three times for each calcium condition. Dose-response values were evaluated in the MO Affinity analysis software and compared between conditions.

***In vitro* analysis of α-syn-induced shedding of sCD163**

Monocyte-derived macrophages (MDMs) were obtained by maturing human monocytes isolated from blood from healthy blood donors as previously described in [13]. Buffy coats (≈ 50mL) from four anonymous healthy donors were obtained from the blood bank at the Department of Clinical Immunology, Aarhus University Hospital (project no. 0094). According to Danish law, the use of anonymized buffy coats does not require specific ethical approval.

Peripheral blood mononuclear cells (PBMCs) were isolated from buffy coats using density gradient centrifugation with a Histopaque-1077 gradient (Sigma-Aldrich, Munich, Germany): Buffy coats were diluted 1:2 in 0.9% NaCl and 25mL diluted blood was carefully layered on 15mL Histopaque-1077. Centrifugation was performed at room temperature, 400g, for 30 min without brake. After centrifugation, the PBMC layer was carefully transferred to a new tube and washed twice in phosphate-buffered saline (PBS) with 2% fetal calf serum (FCS) (Thermofisher Scientific, Waltham, MA) and

1mM EDTA (Merck Milipore, Burlington, MA). Monocytes were isolated using EasySep™ Human Monocyte Isolation Kit (Stemcell Technologies, Vancouver, Canada) according to the manufacturer's instructions. In detail, PBMCs were resuspended in PBS with 2% FCS and 1mM EDTA to a concentration of 5 x $10^7$ cells/mL and incubated with 50µL/mL isolation antibody cocktail and 50µL/mL platelet removal cocktail for 5 min before incubation with 50µL/mL RapidSpheres for 5 min. The PBMCs were subsequently placed in the magnet for 2.5 min and the supernatant, containing the monocytes, was collected and washed once in PBS with1% FCS.

For MDM maturation, the monocytes were resuspended in complete maturation media (RPMI-1640 (ThermoFisher Scientific) with 10% FCS, 100U/100µg/mL penicillin/streptomycin (ThermoFisher Scientific), 10ng/mL macrophage colony-stimulating factor (M-CSF) (Peprotech, Stockholm, Sweden) and 1ng/mL granulocyte-macrophage colony-stimulating factor (GM-CSF) (Peprotech) and seeded in non-treated T-75 flasks. The cells were incubated for 6 days for MDM differentiation in a 5% $CO_2$ container at 37°C. Medium was changes every 2-3 days. After MDM maturation, the medium was removed and the cells were harvested after 15 min incubation in PBS with 0.5% bovine serum albumin, 5mM EDTA, and 4mg/mL lidocaine hydrochloride monohydrate (Sigma Aldrich) using a cell scraper. The MDMs were washed in PBS with 1% FCS and reseeded in RPMI-1640 supplemented with 10% FCS at 1 x $10^6$ cells/mL in 12-well plates with 1mL/well. The MDMs were incubated with 0nM (untreated), 100nM, 1µM, or 5µM monomeric or fibrillar α-syn for 6 or 24 hours or with 100ng/ml LPS as positive control for 1 hour. After incubation, the medium was collected and centrifuged at 500g for 5 min to remove non-adherent cells. The supernatant was transferred to cryovials and placed at -80°C prior to ELISA analysis. Samples were diluted 1:5 for ELISA analysis.

**Statistical analyses**

GraphPad Prism V7, JMP 14, and STATA v15 IC were used for statistical analyses and data plotting. For sex- and group differences as well as correlations, the use of parametric vs. non-parametric tests were determined by testing Gaussian normality distribution with a D'Agostino-Pearson omnibus normality test, followed by unpaired t test (or Mann-Whitney), 1-way ANOVA (or Kruskal-Wallis test) with Tukey's (or Dunn's) multiple comparisons test, and Pearson (or Spearman) correlation coefficient calculations as appropriate.

Simple linear and multiple linear regressions were applied to predict clinical scores of biomarkers for PD patients based on CSF-sCD163 values alone or adjusted for age and/or disease duration. For the analysis of correlations between sCD163 and the Multiplex markers, Bonferroni corrections in the p-values were used.

In Exp#2: ROC analysis of single CSF or serum biomarkers effect on PD diagnosis (PD vs. HC) were estimated using AUC calculations; main effect of PD phenotype stages (UPDRS III, MoCA, Mini Mental (MMSE (O)), or MMSE original plus converted from MoCA (MMSE O+C)) were calculated using linear regression. The main effect of each single biomarkers were plotted as ROC and forest plots. Biomarker contribution to PD prognosis was also investigated using principal component analysis (PCA) for CSF variables. Due to missing values (not all biomarkers were measured in all samples, see above), PCA was not applied for serum biomarkers.

Stepwise forward model strategies were used for a logistic and linear regression with robust variance component estimate for factors affecting PD/HC diagnosis and phenotypical PD scores, respectively. For each diagnosis/score, three stepwise forward prediction models were calculated: Each model is started by selecting the most significant variable with a p value below the entry probability, and then in each step, one additional variable is added and one excluded if the corresponding p values are below entry- and removal probabilities, respectively: M1) probability of entry is 0.1, probability of removal is 0.2; M2) probability of entry is 0.05, probability of removal is 0.1; and M3) probability of entry is 0.1, probability of removal is 0.2, sCD163 forced to be included. Thus, each model is started by selecting the most significant variable, where the following selected variables' significance are non-dependent of the already selected variables. Thus, the numbers of predictors have automatically been reduced

based on different contribution to the diagnose/score. Each variables contribution are provided as log odds ratio or regression slope (column entitled b in **Suppl.Table7**). The three models were compared using AIC and BIC (probabilistic model selection criterion). For the three final models on PD diagnosis and phenotypic scores, a classification table was calculated and Hosmer-Lemeshow goodness of fit was conducted ($\chi 2$). For each model of PD phenotype scores (UPDRS III, MoCA, Mini Mental, or MMSE O+C), the adjusted R squares were calculated and the observed scores were graphed against their fitted value. ROC plots were conducted for the best PD diagnosis models (M3 in both serum and CSF).
For *in vitro* experiments, Gaussian normality distribution was confirmed using a Shapiro-Wilk normality test, Two-way ANOVA was used followed by Tukey's multiple comparisons test.

| Experiment#1 | | PD | HC (CSF & serum) |
|---|---|---|---|
| **Female/male** | Mean | 53/56 | 19/25 |
| ns | Count | 109 | 44 |
| **Age at onset** | Mean ± SD | 61.4 ± 10.7 | - |
| | Min-Max, Count | [30-82], 109 | - |
| **Age at visit** | Mean ± SD | 66.6 ± 9.9 | 64.1± 10.7 |
| ns | Min-Max, Count | [36-84], 109 | [42-81], 44 |
| **Disease duration** | Mean ± SD | 5.2 ± 5.0 | - |
| | Min-Max, Count | [0-22] 109 | - |
| **LEDD** | Mean ± SD | 351.9 ± 372.2 | - |
| | Min-Max, Count | [0-1532], 106 | - |
| **Hoehn & Yahr** | Mean ±S D | 2,2 ± 0.8 | - |
| | Min-Max, Count | [1-4], 105 | - |
| **UPDRS III** | Mean ± SD | 25.6 ± 10.6 | - |
| | Min-Max, Count | [7-66], 96 | - |
| **MoCA** | Mean ± SD | 25.5 ± 4.9 | 26.9 ± 2.2 |
| ns | Min-Max, Count | [11-30], 67 | [23-30], 18 |
| **Mini Mental** | Mean ± SD | 27 ± 3.9 | 28 ± 3.1 |
| ns | Min-Max, Count | [14-30], 100 | [17-30], 30 |
| **MMSE (O+C)** | Mean ± SD | 27.1 ± 3.9 | 28 ± 3.1 |
| ns | Min-Max, Count | [14-30], 105 | [17-30], 30 |
| **CSF Abeta42** | Mean ± SD | 731 ± 266 | 902 ± 307 ** |
| | Min-Max, Count | [118-1435] , 107 | [352-1616], 39 |
| **CSF h-Tau** | Mean ± SD | 244 ± 172 | 273 ± 159 |
| ns | Min-Max, Count | [25-1074], 107 | [112-992], 39 |
| **CSF p-Tau** | Mean ± SD | 41± 19 | 50± 22 * |
| | Min-Max, Count | [15-113], 103 | [22-149], 39 |
| **CSF total α-syn** | Mean ± SD | 561 ± 281 | 496 ± 150 |
| ns | Min-Max, Count | [190-1834], 92 | [226-715], 18 |
| **Albumin CSF/serum x10$^{-3}$** | Mean ± SD | 7,25 ± 3,8 | 6,64 ± 2,1 |
| ns | Min-Max, Count | [2,10-20,58], 66 | [2,72-11,69], 36 |

**Suppl. Table 1 Detailed overview of Exp#1**
Mean, standard derivation (SD), [range] and the number of individuals for whom the data were available at sampling time are shown for: L-dopa equivalent daily dose (LEDD), Unified Parkinson's Disease Rating Scale three (UPDRS III), the Montreal Cognitive Assessment (MoCA) score, the Mini-Mental State Examination (MMSE), MMSE original + converted from MoCA (MMSE O+C). Significance between groups was tested with unpaired t tests: p values with trends are stated, * p< 0.05, ** p < 0.01.

| Experiment#2<br>122 (148) | | PD | HC<br>(CSF & serum) | HC<br>(incl. extra for serum ¤) |
|---|---|---|---|---|
| **Female/male** | Mean | 45 / 61 | 11 / 5 | 20 / 22 |
| | Count | 106 | 16 | 42 |
| **Age at onset** | Mean ± SD | 59.1 ± 10.3 | - | - |
| | Min-Max, Count | [38-76], 106 | - | - |
| **Age at visit** | Mean ± SD | 64.2 ± 10.7 | 56.9 ± 10.9 * | 63.2 ± 11.5 |
| | Min-Max, Count | [41-86],106 | [43-77], 16 | [42-81], 42 |
| **Disease duration** | Mean ± SD | 5.1 ± 3.5 | - | - |
| | Min-Max, Count | [0-17], 106 | - | - |
| **LEDD** | Mean ± SD | 453.7 ± 335.6 | - | - |
| | Min-Max, Count | [0-1574], 106 | - | - |
| **Hoehn and Yahr** | Mean ± SD | 2.0 ± 0.6 | - | - |
| | Min-Max, Count | [1-4], 104 | - | - |
| **UPDRS III** | Mean ± SD | 22.8 ± 11.2 | - | - |
| | Min-Max, Count | [3-66], 101 | - | - |
| **BDI II** | Mean ± SD | 8.3 ± 6.3 | 6.0 ± 1 | 6.8 ± 7.6 |
| | Min-Max, Count | [0-27], 74 | [5-7], 3 | [0-22], 8 |
| **Sniffin sticks 12** | Mean ± SD | 6.7 ± 2.6 | - | 5 |
| | Min-Max, Count | [2-12], 59 | - | 1 |
| **MoCA** | Mean ± SD | 25.8 ± 3.6 | 27.7 ± 4 | 27.0 ± 2.4 |
| | Count | [11-30], 93 | [23-30], 3 | [23-30], 14 |
| **MMSE (O)** | Mean ± SD | 28.1 ± 2.3 | 28.3 ± 2.9 | 28.0 ± 3.4 |
| | Min-Max, Count | [17-30], 89 | [23-30], 6 | [17-30], 24 |
| **MMSE (O+C)** | Mean ± SD | 28.1 ± 2.3 | 28.3 ± 2.9 | 28.0 ± 3.4 |
| | Min-Max, Count | [17-30], 101 | [23-30], 6 | [17-30], 24 |
| **CSF Abeta42** | Mean ± SD | 701 ± 246.9 | 787 ± 248.3 | - |
| | Min-Max, Count | [256-1312], 104 | [352-1215], 16 | - |
| **CSF h-Tau** | Mean ± SD | 221.7 ± 109.5 | 233.8 ± 213.3 | - |
| | Min-Max, Count | [57-737], 104 | [129-992], 16 | - |
| **CSF p-Tau** | Mean ± SD | 38.6 ± 14.6 | 42.3 ± 29.7 | - |
| | Min-Max, Count | [15-97], 103 | [18-149], 16 | - |
| **CSF total α-syn** | Mean ± SD | 631 ± 274.7 | 531 ± 101.6 | - |
| | Min-Max, Count | [200-1486], 71 | [289-664], 13 | - |

**Suppl. Table 2 Detailed overview of exp#2**
Mean, standard derivation (SD), [range] and the number of individuals for whom the data were available at sampling time are shown for: L-dopa equivalent daily dose (LEDD), Unified Parkinson's Disease Rating Scale three (UPDRS III), Beck Depression Inventory II (BDI II), the Montreal Cognitive Assessment (MoCA) score, the Mini-Mental State Examination (MMSE) original score (O), MMSE original + converted from MoCA (MMSE O+C). ¤: Extra 26 serum controls from Exp#1 were added, since measurements of nine cytokines were unaffected by an extra freezing/thaw cycle, for details see **Suppl.Table1**. Significant differences were tested between PD and HC groups with unpaired t tests: * p 0.0124.

| Method | Soluble molecule | Serum Measured | Serum <LLOD | CSF Measured | CSF <LLOD |
|---|---|---|---|---|---|
| ELISA | sCD163 | yes | | yes | |
| ELISA | a-syn | no | | yes | |
| ELISA | h-Tau | no | | yes | |
| ELISA | p-Tau | no | | yes | |
| ELISA | AB42 | no | | yes | |
| MSD VIP2 | SAA | yes | | yes | |
| MSD VIP2 | CRP | yes ¤ | | yes | |
| MSD VIP2 | VCAM-1 | yes | | yes | |
| MSD VIP2 | ICAM-1 | yes | | yes | |
| MSD AP1 | VEGF-A | yes | | yes | |
| MSD AP1 | VEGF-C | yes ¤ | | yes | yes |
| MSD AP1 | VEGF-D | yes | | yes | (3) |
| MSD AP1 | Tie-2 | yes | | yes | yes |
| MSD AP1 | Flt-1 | yes ¤ | | yes | |
| MSD AP1 | PIGF | yes ¤ | | yes | |
| MSD AP1 | bFGF | yes ¤ | | yes | yes |
| MSD CyP1 | GM-CSF | yes | yes | yes | yes |
| MSD CyP1 | IL-1a | yes | yes | yes | yes |
| MSD CyP1 | IL-5 | yes | yes | yes | yes |
| MSD CyP1 | IL-7 | yes ¤ | | yes | |
| MSD CyP1 | IL-12/IL-23p40 | yes ¤ | | yes | |
| MSD CyP1 | IL-15 | yes ¤ | | yes | |
| MSD CyP1 | IL-16 | yes ¤ | | yes | yes |
| MSD CyP1 | IL-17A | yes | yes | yes | yes |
| MSD CyP1 | TNFb | yes | yes | yes | yes |
| MSD CyP1 | (VEGF-A) | NA | NA | NA | NA |
| MSD ChP1 | Eotaxin | yes | | yes | yes |
| MSD ChP1 | MIP-1a (CCL3) | yes | yes | yes | yes |
| MSD ChP1 | Eotaxin3 | yes | (2) | yes | yes |
| MSD ChP1 | TARC | yes | | yes | yes |
| MSD ChP1 | IP-10 (CXCL10) | yes | | yes | |
| MSD ChP1 | MIP-1b (CCL4) | yes | | yes | |
| MSD ChP1 | (IL-8HA) | NA | NA | NA | NA |
| MSD ChP1 | MCP-1 (CCL2) | yes | | yes | |
| MSD ChP1 | MDC (CCL22) | yes | | yes | yes |
| MSD ChP1 | MCP-4 (CCL13) | yes | | yes | yes |
| MSD PP1 | IFNg | yes | yes | yes | yes |
| MSD PP1 | IL-1b | yes | yes | yes | yes |
| MSD PP1 | IL-2 | yes | yes | yes | yes |
| MSD PP1 | IL-4 | yes | yes | yes | yes |
| MSD PP1 | Il-6 | yes | (1) | yes | |
| MSD PP1 | IL-8 | yes | | yes | |
| MSD PP1 | IL-10 | yes | | yes | yes |
| MSD PP1 | IL12p70 | yes | yes | yes | yes |
| MSD PP1 | IL-13 | yes | yes | yes | yes |
| MSD PP1 | TNFa | yes | | yes | yes |

**Suppl. Table 3 Assay overview for Exp#2**

Mesoscale 40-plex (MSD) panels: Vascular injury panel 2 (VIP2), Angiogenesis panel 1 (AP1), Cytokine panel 1 (CyP1), Chemokine panel 1 (ChP1), Pro-inflammatory panel 1 (PP1). NA: Not applied. ¤: Extra 26 serum controls from Exp#1 were added, since measurements were unaffected by extra freezing/thaw cycle. Numerous samples with values below lower limit of detection (>LLOD), thus analysis was excluded; if only a few (n in brackets) samples had values <LLOD, those measurements were replaced with a value equal to LLOD/2 and include in analysis according to MSD guidelines.

| Linear regression model | Dependent variable | Independent variable | Estimate | Std Error | t Ratio | Prob>\|t\| | Lower 95% | Upper 95% | Rsqu. |
|---|---|---|---|---|---|---|---|---|---|
| **Simple** | α-syn | CSF sCD163 | 2241.60 | 687.74 | 3.26 | **0.0015** | 878.37 | 3604.83 | 0.089 |
| **Simple** | α-syn | age at visit | 6.44 | 2.43 | 2.65 | **0.0094** | 1.61 | 11.26 | 0.065 |
| **Simple** | α-syn | disease duration | 4.14 | 5.90 | 0.7 | 0.4845 | -7.58 | 15.87 | 0.005 |
| **Multiple** | α-syn | disease duration | -1.98 | 6.10 | -0.33 | 0.7453 | -14.10 | 10.13 | 0.142 |
| | | age at visit | 5.34 | 3.03 | 1.76 | 0.0816 | -0.68 | 11.36 | |
| | | CSF sCD163 mg/ml | 2088.96 | 898.71 | 2.32 | **0.0224** | 302.97 | 3874.95 | |
| **Multiple** | α-syn | age at visit | 4.27 | 2.53 | 1.69 | 0.094 | -0.73 | 9.28 | 0.113 |
| | | CSF sCD163 mg/ml | 1823.57 | 725.41 | 2.51 | **0.0134** | 385.53 | 3261.61 | |
| **Multiple** | α-syn | disease duration | -3.68 | 6.09 | -0.60 | 0.5469 | -15.78 | 8.42 | 0.112 |
| | | CSF sCD163 mg/ml | 2728.20 | 831.85 | 3.28 | **0.0015** | 1075.33 | 4381.08 | |
| **Simple** | p-Tau | CSF sCD163 mg/ml | 204.13 | 51.62 | 3.95 | **0.0001** | 102.07 | 306.18 | 0.1004 |
| **Simple** | p-Tau | age at visit | 0.54 | 0.17 | 3.12 | **0.0022** | 0.20 | 0.89 | 0.065 |
| **Simple** | p-Tau | disease duration | 0.59 | 0.38 | 1.56 | 0.1216 | -0.16 | 1.35 | 0.02 |
| **Multiple** | p-Tau | disease duration | 0.24 | 0.37 | 0.63 | 0.5277 | -0.50 | 0.97 | 0.213 |
| | | age at visit | 0.61 | 0.19 | 3.17 | **0.0020** | 0.23 | 0.99 | |
| | | CSF sCD163 mg/ml | 136.42 | 56.66 | 2.41 | **0.0179** | 23.99 | 248.84 | |
| **Multiple** | p-Tau | age at visit | 0.37 | 0.18 | 2.09 | **0.0381** | 0.02 | 0.72 | 0.127 |
| | | CSF sCD163 mg/ml | 169.61 | 53.61 | 3.16 | **0.0019** | 63.62 | 275.60 | |
| **Multiple** | p-Tau | disease duration | 0.10 | 0.39 | 0.26 | 0.7954 | -0.66 | 0.87 | 0.133 |
| | | CSF sCD163 mg/ml | 198.31 | 55.54 | 3.57 | **0.0005** | 88.11 | 308.50 | |
| **Simple** | h-Tau | CSF sCD163 mg/ml | 1761.95 | 401.38 | 4.39 | **<.0001** | 968.59 | 2555.31 | 0.118 |
| **Simple** | h-Tau | age at visit | 5.34 | 1.40 | 3.82 | **0.0002** | 2.58 | 8.11 | 0.092 |
| **Simple** | h-Tau | disease duration | 5.93 | 3.27 | 1.81 | 0.073 | -0.56 | 12.42 | 0.03 |
| **Multiple** | h-Tau | disease duration | 2.00 | 3.31 | 0.61 | 0.5463 | -4.55 | 8.56 | 0.207 |
| | | age at visit | 5.31 | 1.73 | 3.07 | **0.0028** | 1.88 | 8.74 | |
| | | CSF sCD163 mg/ml | 1150.21 | 499.93 | 2.3 | **0.0234** | 158.72 | 2141.70 | |
| **Multiple** | h-Tau | age at visit | 3.75 | 1.43 | 2.62 | **0.0098** | 0.92 | 6.58 | 0.15 |
| | | CSF sCD163 mg/ml | 1400.11 | 417.05 | 3.36 | **0.0010** | 575.73 | 2224.49 | |
| **Multiple** | h-Tau | disease duration | 0.91 | 3.42 | 0.26 | 0.7916 | -5.87 | 7.68 | 0.134 |
| | | CSF sCD163 mg/ml | 1712.45 | 483.55 | 3.54 | **0.0006** | 753.55 | 2671.34 | |
| **Linear regression model** | **Dependent variable** | **Independent variable** | **Estimate** | **Std Error** | **t Ratio** | **Prob>\|t\|** | **Lower 95%** | **Upper 95%** | **Rsqu.** |
| **Simple** | p-Tau | Serum-sCD163 | 6.254 | 2.2983 | 2.72 | **0.0073** | 1.7099 | 10.798 | 0.0505 |
| **Multiple** | p-Tau | age at visit | 0.6743 | 0.1831 | 3.68 | **0.0004** | 0.311 | 1.0376 | 0.220 |
| | | Disease duration | 0.4139 | 0.3488 | 1.19 | 0.2382 | -0.278208 | 1.106 | |
| | | Serum-sCD163 | 5.79 | 2.2366 | 2.59 | **0.0111** | 1.352 | 10.228 | |
| **Multiple** | p-Tau | age at visit | 0.4648 | 0.1764 | 2.63 | **0.0094** | 0.116 | 0.8135 | 0.096 |
| | | Serum-sCD163 | 5.2694 | 2.2815 | 2.31 | **0.0224** | 0.7582 | 9.7805 | |
| **Simple** | h-Tau | Serum-sCD163 | 60.467 | 18.579 | 3.25 | **0.0014** | 23.742 | 97.192 | 0.068 |
| **Multiple** | h-Tau | age at visit | 6.1021 | 1.6229 | 3.76 | **0.0003** | 2.8834 | 9.3208 | 0.214 |
| | | Disease duration | 4.2186 | 3.0067 | 1.4 | 0.1636 | -1.7445 | 10.182 | |
| | | Serum-sCD163 | 49.458 | 19.642 | 2.52 | **0.0133** | 10.502 | 88.413 | |
| **Multiple** | h-Tau | age at visit | 4.7271 | 1.4038 | 3.37 | **0.0010** | 1.952 | 7.5021 | 0.137 |
| | | Serum-sCD163 | 51.802 | 18.125 | 2.86 | **0.0049** | 15.972 | 87.632 | |
| **Simple** | α-syn | Serum-sCD163 | 135.21 | 31.597 | 4.28 | **<.0001** | 72.584 | 197.84 | 0.144 |
| **Multiple** | α-syn | age at visit | 6.4793 | 2.7031 | 2.4 | **0.0186** | 1.1074 | 11.851 | 0.209 |
| | | Disease duration | 1.5195 | 5.362 | 0.28 | 0.7776 | -9.136341 | 12.175 | |
| | | Serum-sCD163 | 125.89 | 34.56 | 3.64 | **0.0005** | 57.213 | 194.57 | |
| **Multiple** | α-syn | age at visit | 4.8907 | 2.3193 | 2.11 | **0.0373** | 0.293 | 9.4884 | 0.179 |
| | | Serum-sCD163 | 123.92 | 31.562 | 3.93 | **0.0002** | 61.347 | 186.48 | |

**Suppl. Table 4 Simple linear and multiple linear regression were calculated to predict the different neuronal CSF markers for PD patients based on CSF-sCD163 (top) or serum sCD163 (bottom) values alone or adjusted for age and/or disease duration for Exp#1** Estimate of slope (β)/model coefficients with standard (std) errors, the t ratio of the estimate to the std error (t Ratio), the p values (Prob>ItI) (bold if significant), the 95% lower and upper confidence limits for the parameter estimate and the Rsquare (Rsqu) values are shown for simple linear regressions or multiple regression analyses.

| Linear regression model | Dependent variable | Independent variable | Estimate | Std Error | t Ratio | Prob>\|t\| | Lower 95% | Upper 95% |
|---|---|---|---|---|---|---|---|---|
| Simple | MMSE (O+C) | **CD163 mg/l** | **-29.98** | 6.68 | -4.49 | **<.0001** | -43.24 | -16.72 |
| Simple | MMSE (O+C) | age at visit | -0.09 | 0.02 | -4.6 | **<.0001** | -0.13 | -0.05 |
| Simple | MMSE (O+C) | age at onset | -0.08 | 0.02 | -3.79 | **0.0003** | -0.12 | -0.04 |
| Simple | MMSE (O+C) | disease duration | -0.16 | 0.06 | -2.42 | **0.02** | -0.28 | -0.03 |
| Multiple | MMSE (O+C) | age at onset | -0.044 | 0.02 | -1.81 | **0.0733** | -0.09 | 0 |
| | | **CD163 mg/l** | **-22.58** | 7.77 | -2.91 | **0.0045** | -37.99 | -7.16 |
| Multiple | MMSE (O+C) | disease duration | -0.1 | 0.06 | -1.58 | 0.117 | -0.22 | 0.02 |
| | | **CD163 mg/l** | -27.45 | 6.82 | -4.02 | **0.0001** | -40.99 | -13.92 |
| Multiple | MMSE (O+C) | age at visit | -0.06 | 0.02 | -2.49 | **0.0146** | -0.11 | -0.01 |
| | | **CD163 mg/l** | **-18.36** | 8.01 | -2.29 | **0.0242** | -34.27 | -2.45 |
| Multiple | MMSE (O+C) | age at onset | -0.05 | 0.02 | -2.23 | **0.028** | -0.1 | -0.01 |
| | | disease duration | -0.13 | 0.06 | -2.05 | **0.0433** | -0.25 | 0 |
| | | **CD163 mg/l** | **-17.52** | 8.03 | -2.18 | **0.0316** | -33.47 | -1.58 |
| Multiple | MMSE (O+C) | age at visit | -0.05 | 0.02 | -2.23 | **0.028** | -0.1 | -0.01 |
| | | disease duration | -0.07 | 0.06 | -1.17 | 0.2439 | -0.19 | 0.05 |
| | | **CD163 mg/l** | **-17.52** | 8.03 | -2.18 | **0.0316** | -33.47 | -1.58 |
| Multiple | MMSE (O+C) | age at onset | 0.07 | 0.06 | 1.17 | 0.2439 | -0.05 | 0.19 |
| | | age at visit | -0.13 | 0.06 | -2.05 | **0.0433** | -0.25 | 0 |
| | | **CD163 mg/l** | **-17.52** | 8.04 | -2.18 | **0.0316** | -33.47 | -1.58 |

**Suppl. Table 5 Simple linear and multiple linear regression were calculated to predict MMSE (O+C) scores for PD patients based on CSF-sCD163 values alone or adjusted for age and/or disease duration for Exp#2**

Estimate of slope (β)/model coefficients with standard (std) errors, the t ratio of the estimate to the std error (t Ratio), the p values (Prob>ItI) (significant values are marked with bold), and the 95% lower and upper confidence limits for the parameter estimate are shown for simple linear regressions or multiple regression analyses. The Mini-Mental State Exam MMSE original plus converted from Montreal Cognitive Assessment (MoCA) (O+C) score.

| Sample | Clinical prediction | Model | N | ll0 | ll | df | AIC | BIC | Selected model | Goodness of fit (R) |
|---|---|---|---|---|---|---|---|---|---|---|
| serum | PD/HC | M1 | 122 | -47.40 | -34.09 | 6 | 80.19 | 97.01 | | (χ2) 0.82 Cc 90.2% |
| serum | PD/HC | M2 | 122 | -47.40 | -37.17 | 3 | 80.34 | 88.75 | | (χ2) 0.21 Cc 86.9% |
| serum | PD/HC | M3 | 122 | -47.40 | -30.96 | 8 | 77.91 | 100.35 | x | (χ2) 0.76 Cc 89.3% |
| serum | UPDRS III | M1 | 101 | -386.55 | -368.84 | 10 | 757.68 | 783.84 | x | 0.23 |
| serum | UPDRS III | M2 | 101 | -386.55 | -376.50 | 5 | 763.00 | 776.08 | | 0.15 |
| serum | UPDRS III | M3 | 101 | -386.55 | -368.45 | 11 | 758.90 | 787.67 | | 0.22 |
| serum | MoCA | M1 | 93 | -251.78 | -233.01 | 6 | 478.01 | 493.21 | | 0.29 |
| serum | MoCA | M2 | 93 | -251.78 | -237.92 | 3 | 481.84 | 489.44 | | 0.24 |
| serum | MoCA | M3 | 93 | -251.78 | -232.92 | 7 | 479.85 | 497.58 | x | 0.29 |
| serum | MiniMental | M1 | 89 | -200.30 | -184.80 | 5 | 379.60 | 392.05 | x | 0.26 |
| serum | MiniMental | M2 | 89 | -200.30 | -187.61 | 4 | 383.21 | 393.17 | | 0.22 |
| serum | MiniMental | M3 | 89 | -200.30 | -184.59 | 6 | 381.18 | 396.11 | | 0.26 |
| serum | MMSE (O+C) | M1 | 101 | -226.94 | -209.42 | 7 | 432.84 | 451.14 | x | 0.25 |
| serum | MMSE (O+C) | M2 | 101 | -226.94 | -216.14 | 4 | 440.27 | 450.73 | | 0.17 |
| serum | MMSE (O+C) | M3 | 101 | -226.94 | -209.61 | 9 | 437.22 | 460.76 | | 0.23 |
| CSF | PD/HC | M1 | 83 | -36.02 | -21.40 | 6 | 54.79 | 69.31 | | (χ2) 0.86 Cc 86.75 |
| CSF | PD/HC | M2 | 83 | -36.02 | -28.56 | 3 | 63.12 | 70.38 | | (χ2) 0.26 Cc 86.8% |
| CSF | PD/HC | M3 | 83 | -36.02 | -15.35 | 8 | 46.70 | 66.05 | x | (χ2) 0.80 Cc 90.4% |
| CSF | UPDRS III | M1 | 67 | -257.66 | -255.40 | 2 | 514.80 | 519.21 | x | 0.05 |
| CSF | UPDRS III | M2 | 67 | -257.66 | -255.40 | 2 | 514.80 | 519.21 | | 0.05 |
| CSF | UPDRS III | M3 | 67 | -257.66 | -255.17 | 3 | 516.34 | 522.96 | | 0.04 |
| CSF | MoCA | M1 | 63 | -159.10 | -143.84 | 6 | 299.67 | 312.53 | | 0.33 |
| CSF | MoCA | M2 | 63 | -159.10 | -146.27 | 4 | 300.55 | 309.12 | | 0.30 |
| CSF | MoCA | M3 | 63 | -159.10 | -144.62 | 5 | 299.24 | 309.96 | x | 0.32 |
| CSF | MiniMental | M1 | 62 | -128.92 | -125.47 | 3 | 256.94 | 263.32 | | 0.07 |
| CSF | MiniMental | M2 | 62 | -128.92 | -127.86 | 2 | 259.71 | 263.96 | | 0.02 |
| CSF | MiniMental | M3 | 62 | -128.92 | -125.53 | 3 | 257.05 | 263.43 | x | 0.07 |
| CSF | MMSE (O+C) | M1 | 67 | -138.80 | -134.91 | 3 | 275.83 | 282.44 | | 0.08 |
| CSF | MMSE (O+C) | M2 | 67 | -138.80 | -134.91 | 3 | 275.83 | 282.44 | | 0.08 |
| CSF | MMSE (O+C) | M3 | 67 | -138.80 | -134.12 | 4 | 276.23 | 285.05 | x | 0.09 |

**Suppl. Table 6 Stepwise forward models overview (Exp#2):**
Stepwise forward model strategies were used for a logistic regression with robust variance component estimation for factors affecting PD/HC diagnosis; and linear regression with robust variance component estimation for factors with effect on PD progression scores (UPDRS III, MoCA, Mini Mental original score (MMSE (O), or MMSE original score plus converted from MoCA (MMSE (O+C)) in Exp#2. For each diagnosis/score, three models were calculated: M1) probability of entry is 0.1, probability of removal is 0.2; M2) probability of entry is 0.05, probability of removal is 0.1; M3) probability of entry is 0.1, probability of removal is 0.2, CD163 forced to be included. The models are compared using AIC and BIC and goodness of fit for: Rsquare for the linear regression models and chi2 (χ2) for the logistic models (marked with gray). Best models (x) for each prediction score are plotted in **Fig.4M-N & Suppl.Fig.7**. Details of models are described in **Suppl.Table 7**.

| Sample | Clinical prediction | Biomarker | M1, b | 95% lower CI | 95% upper CI | P | M2, b | 95% lower CI | 95% upper CI | P | M3, b | 95% lower CI | 95% upper CI | P |
|---|---|---|---|---|---|---|---|---|---|---|---|---|---|---|
| serum | PD/HC | VCAM-1 | 0.151 | 0.053 | 0.248 | 0.00 | 0.129 | 0.044 | 0.213 | 0.00 | 0.145 | 0.032 | 0.259 | 0.01 |
| serum | PD/HC | Flt-1 | -0.812 | -1.620 | -0.005 | 0.05 | -1009 | -1838 | -0.180 | 0.02 | -0.992 | -1.821 | -0.163 | 0.02 |
| serum | PD/HC | VEGF-D | -0.211 | -0.428 | 0.006 | 0.06 | | | | | -0.226 | -0.475 | 0.023 | 0.07 |
| serum | PD/HC | IP-10 | -0.267 | -0.553 | 0.019 | 0.07 | | | | | | | | |
| serum | PD/HC | IL-15 | 1.092 | -0.146 | 2.331 | 0.08 | | | | | 1.777 | 0.156 | 3.398 | 0.03 |
| serum | PD/HC | **sCD163** | | | | | | | | | 0.829 | -0.591 | 2.248 | 0.25 |
| serum | PD/HC | IL-10 | | | | | | | | | -3.041 | -5.595 | -0.488 | 0.02 |
| serum | PD/HC | MIP-1β | | | | | | | | | 1.389 | -0.264 | 3.042 | 0.10 |
| serum | UPDRS III | VEGF-A | -1.072 | -1.603 | -0.542 | 0.00 | -1050 | -1.536 | -0.565 | 0.00 | -1.100 | -1.638 | -0.562 | 0.00 |
| serum | UPDRS III | PIGF | 1.994 | 0.845 | 3.142 | 0.00 | 1678 | 0.602 | 2.753 | 0.00 | 2.052 | 0.879 | 3.226 | 0.00 |
| serum | UPDRS III | SAA | -0.114 | -0.170 | -0.059 | 0.00 | -0.094 | -0.170 | -0.018 | 0.02 | -0.117 | -0.172 | -0.062 | 0.00 |
| serum | UPDRS III | IL-8 | 0.107 | 0.010 | 0.204 | 0.03 | 0.112 | 0.007 | 0.216 | 0.04 | 0.121 | 0.025 | 0.217 | 0.01 |
| serum | UPDRS III | TARC | 1.851 | 0.391 | 3.311 | 0.01 | | | | | 1.845 | 0.383 | 3.306 | 0.01 |
| serum | UPDRS III | IL-16 | -2.558 | -4.418 | -0.698 | 0.01 | | | | | -2.624 | -4.525 | -0.723 | 0.01 |
| serum | UPDRS III | MCP-1 | -2.232 | -3.863 | -0.601 | 0.01 | | | | | -2.345 | -4.031 | -0.659 | 0.01 |
| serum | UPDRS III | MCP-4 | 4.616 | 0.882 | 8.351 | 0.02 | | | | | 4.801 | 1.025 | 8.578 | 0.01 |
| serum | UPDRS III | CRP | 0.354 | -0.045 | 0.752 | 0.08 | | | | | 0.380 | -0.039 | 0.799 | 0.08 |
| serum | UPDRS III | **sCD163** | | | | | | | | | -1.146 | -3.102 | 0.809 | 0.25 |
| serum | MoCA | IL-15 | -1.645 | -2.888 | -0.402 | 0.01 | -2212 | -3.502 | -0.921 | 0.00 | -1.631 | -2.861 | -0.402 | 0.01 |
| serum | MoCA | CRP | -0.266 | -0.455 | -0.076 | 0.01 | -0.266 | -0.484 | -0.049 | 0.02 | -0.268 | -0.457 | -0.079 | 0.01 |
| serum | MoCA | MCP-1 | 0.829 | 0.140 | 1.518 | 0.02 | | | | | 0.834 | 0.139 | 1.530 | 0.02 |
| serum | MoCA | PIGF | -0.416 | -0.748 | -0.083 | 0.01 | | | | | -0.418 | -0.750 | -0.085 | 0.01 |
| serum | MoCA | IL-7 | -0.118 | -0.237 | 0.002 | 0.05 | | | | | -0.119 | -0.239 | 0.002 | 0.05 |
| serum | MoCA | **sCD163** | | | | | | | | | 0.170 | -0.636 | 0.976 | 0.68 |
| serum | Mini Mental | Eotaxin-3 | 0.028 | 0.009 | 0.047 | 0.01 | 0.021 | 0.006 | 0.036 | 0.01 | 0.029 | 0.009 | 0.049 | 0.01 |
| serum | Mini Mental | ICAM-1 | 0.049 | 0.024 | 0.073 | 0.00 | 0.038 | 0.015 | 0.062 | 0.00 | 0.047 | 0.024 | 0.069 | 0.00 |
| serum | Mini Mental | CRP | -0.192 | -0.355 | -0.028 | 0.02 | -0.199 | -0.362 | -0.035 | 0.02 | -0.192 | -0.354 | -0.030 | 0.02 |
| serum | Mini Mental | Eotaxin | -0.559 | -1.151 | 0.033 | 0.06 | | | | | -0.568 | -1.158 | 0.022 | 0.06 |
| serum | Mini Mental | **sCD163** | | | | | | | | | 0.186 | -0.269 | 0.641 | 0.42 |
| serum | MMSE (O+C) | Eotaxin | -0.539 | -1.047 | -0.030 | 0.04 | | | | | -0.534 | -1.044 | -0.024 | 0.04 |
| serum | MMSE (O+C) | ICAM-1 | 0.048 | 0.008 | 0.088 | 0.02 | 0.033 | 0.010 | 0.057 | 0.01 | 0.023 | 0.001 | 0.046 | 0.04 |
| serum | MMSE (O+C) | Eotaxin-3 | 0.023 | 0.008 | 0.038 | 0.00 | 0.018 | 0.003 | 0.033 | 0.02 | 0.024 | 0.009 | 0.039 | 0.00 |
| serum | MMSE (O+C) | CRP | -0.174 | -0.323 | -0.026 | 0.02 | -0.185 | -0.350 | -0.019 | 0.03 | -0.155 | -0.322 | 0.011 | 0.07 |
| serum | MMSE (O+C) | VEGF-D | -0.154 | -0.292 | -0.016 | 0.03 | | | | | | | | |
| serum | MMSE (O+C) | Tie-2 | 0.061 | -0.003 | 0.124 | 0.06 | | | | | | | | |
| serum | MMSE (O+C) | **sCD163** | | | | | | | | | 0.053 | -0.395 | 0.500 | 0.82 |
| serum | MMSE (O+C) | IL-8 | | | | | | | | | -0.038 | -0.071 | -0.005 | 0.02 |
| serum | MMSE (O+C) | TNF-α | | | | | | | | | 1.071 | -0.004 | 2.146 | 0.05 |
| serum | MMSE (O+C) | SAA | | | | | | | | | -0.012 | -0.026 | 0.001 | 0.08 |
| CSF | PD/HC | h-Tau | 0.014 | 0.004 | 0.024 | 0.01 | 0.017 | 0.007 | 0.026 | 0.00 | 0.017 | -0.002 | 0.036 | 0.08 |
| CSF | PD/HC | Abeta42 | -0.005 | -0.008 | -0.002 | 0.00 | -0.004 | -0.007 | -0.002 | 0.00 | -0.006 | -0.010 | -0.002 | 0.00 |
| CSF | PD/HC | SAA | 0.003 | 0.001 | 0.006 | 0.01 | | | | | 0.003 | -0.000 | 0.006 | 0.10 |
| CSF | PD/HC | VEGF-A | -0.046 | -0.088 | -0.005 | 0.03 | | | | | -0.093 | -0.157 | -0.030 | 0.00 |
| CSF | PD/HC | IL-6 | 0.203 | 0.025 | 0.381 | 0.03 | | | | | | | | |
| CSF | PD/HC | **sCD163** | | | | | | | | | 0.075 | 0.006 | 0.144 | 0.03 |
| CSF | PD/HC | VCAM-1 | | | | | | | | | 0.001 | 0.000 | 0.002 | 0.01 |
| CSF | PD/HC | Flt-1 | | | | | | | | | -0.216 | -0.377 | -0.056 | 0.01 |
| CSF | UPDRS III | Flt-1 | -0.202 | -0.400 | -0.004 | 0.05 | -0.202 | -0.400 | -0.004 | 0.05 | -0.248 | -0.480 | -0.015 | 0.04 |
| CSF | UPDRS III | **sCD163** | | | | | | | | | 0.036 | -0.066 | 0.138 | 0.48 |
| CSF | MoCA | p-Tau | -0.071 | -0.114 | -0.028 | 0.00 | -0.083 | -0.128 | -0.038 | 0.00 | -0.048 | -0.102 | 0.006 | 0.08 |
| CSF | MoCA | IL-7 | -0.397 | -0.615 | -0.179 | 0.00 | -0.412 | -0.646 | -0.178 | 0.00 | -0.412 | -0.626 | -0.197 | 0.00 |
| CSF | MoCA | IL-8 | 0.090 | 0.023 | 0.157 | 0.01 | 0.081 | 0.013 | 0.148 | 0.02 | 0.089 | 0.024 | 0.154 | 0.01 |
| CSF | MoCA | VCAM-1 | -0.001 | -0.001 | -0.000 | 0.02 | | | | | | | | |
| CSF | MoCA | ICAM-1 | 0.001 | -0.000 | 0.003 | 0.08 | | | | | | | | |
| CSF | MoCA | **sCD163** | | | | | | | | | -0.025 | -0.055 | 0.005 | 0.10 |
| CSF | Mini Mental | IL-12/IL-23p40 | 0.028 | 0.011 | 0.044 | 0.00 | 0.017 | 0.002 | 0.032 | 0.02 | 0.025 | 0.008 | 0.042 | 0.00 |
| CSF | Mini Mental | p-Tau | -0.036 | -0.072 | 0.000 | 0.05 | | | | | | | | |
| CSF | Mini Mental | **sCD163** | | | | | | | | | -0.017 | -0.035 | 0.000 | 0.05 |
| CSF | MMSE (O+C) | IL-12/IL-23p40 | 0.025 | 0.007 | 0.043 | 0.01 | 0.025 | 0.007 | 0.043 | 0.01 | 0.024 | 0.007 | 0.041 | 0.01 |
| CSF | MMSE (O+C) | IL-15 | -0.077 | -0.142 | -0.013 | 0.02 | -0.077 | -0.142 | -0.013 | 0.02 | | | | |
| CSF | MMSE (O+C) | **sCD163** | | | | | | | | | -0.018 | -0.034 | -0.001 | 0.04 |
| CSF | MMSE (O+C) | IL-6 | | | | | | | | | -0.078 | -0.169 | 0.012 | 0.09 |

**Suppl. Table 7 Stepwise forward models' details (Exp#2)**

Variables included in each stepwise forward model (M1-3) are listed; non-selected models/variables are shaded with gray. M1) probability of entry is 0.1, probability of removal is 0.2; M2) probability of entry is 0.05, probability of removal is 0.1;

M3) probability of entry is 0.1, probability of removal is 0.2, CD163 forced to be included. The contribution of each variable are estimated as log odds ratio or regression slope for the respective calculations (b). 0.00 equals all p values <0.01. Selected models are plotted in **Fig. 4M-N & Suppl.Fig.7**.

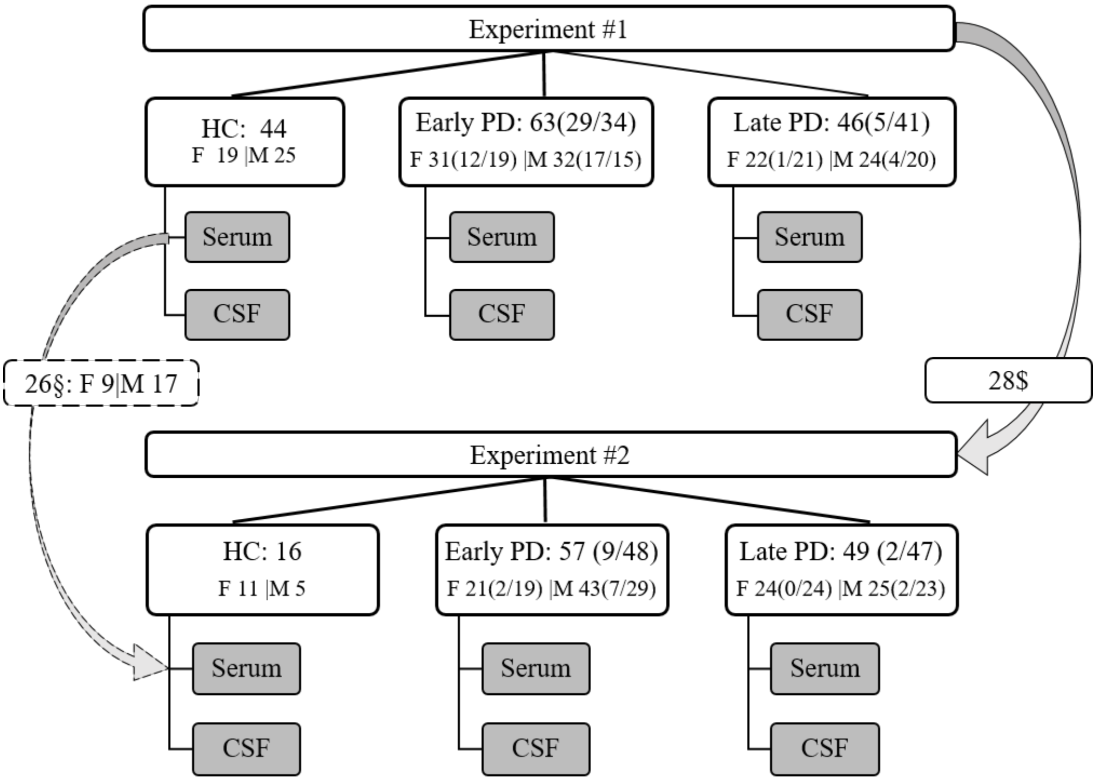

**Suppl. Fig. 1 Overview of individuals included in the two experiments**
Cross-sectional biobank samples of serum and CSF in two experiments (Exp) with a two-year time gap. Exp#1 consists of a total of 153 individuals, 44 healthy controls (HCs) and 109 Parkinson's disease (PD) patients with early [<5 years since diagnosis] or late [≥5 years since diagnosis] stage disease. Treatment status of patients are shown in brackets; untreated (UT)/treated (T). More details of participant in Exp#1 are found **in Supp.Table1**. Exp#2 includes a total of 122 individuals, 16 HCs and 106 PD patients with early or late status. **$**: Twenty-eight individuals from Exp#1 had new aliquots of the biological samples included and analyzed in Exp#2 (4 HC females (F), 4 F early PD (1UT/3T), 3 F late PD (1UT/2T), 10 male (M) early PD (7UT/ 3T), 7 M late PD (2UT/5T)). More details of Exp#2 are found in **Supp.Table2** **§**: Twenty-six serum samples from controls (9 F, 17 M) from Exp#1 were reused for in Exp#2 for analyzing nine cytokines, which were found insensitive for repeated thawing using 40-plex mesoscale. See **Suppl.Table3** for an assay overview for Exp#2.

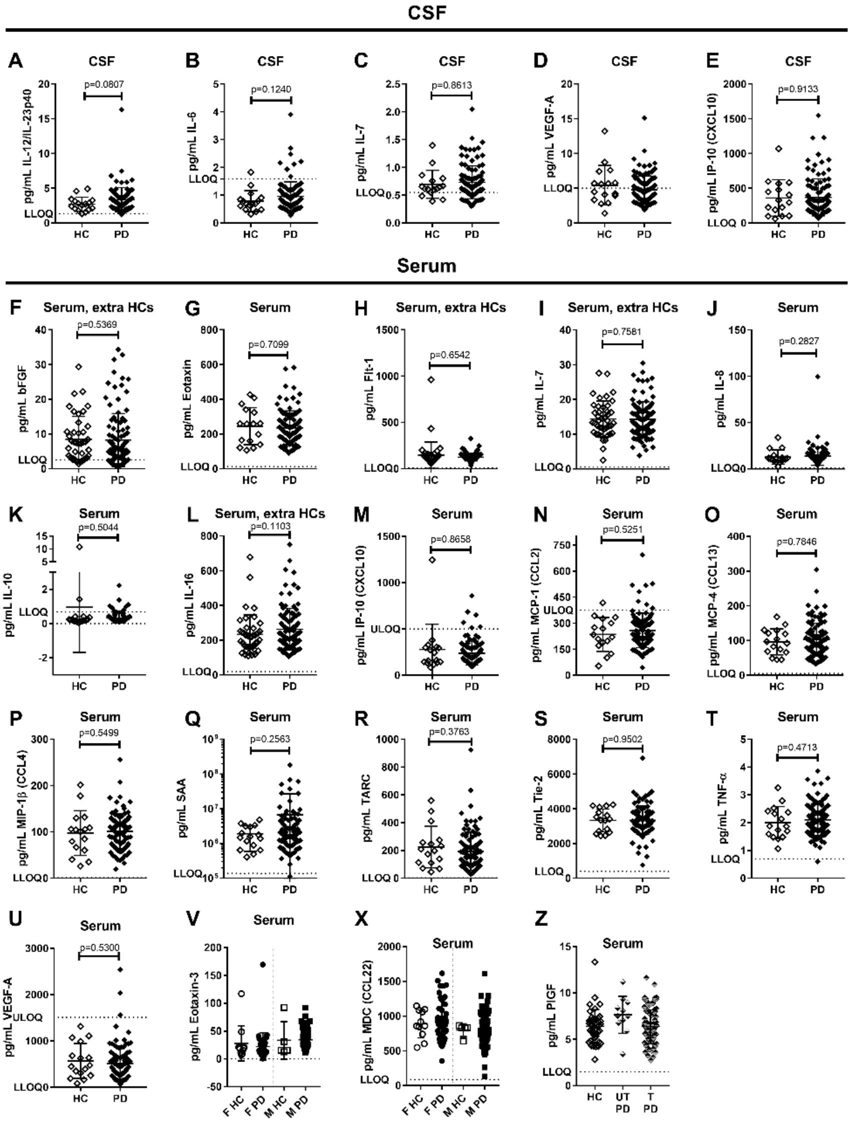

**Suppl. Fig. 2 Additional CSF and serum biomarkers (Exp#2);**

Sixteen different CSF biomarkers and twenty seven serum biomarkers from the 40-plex mesoscale assay had values >LLOD (lower limit of detection) and were tested for differences between healthy control (HC) and Parkinson's disease (PD) patient groups; or with respect to progression from early to late; or with respect to treatment status with or without L-dopa. CSF biomarkers **A-E)** and serum biomarkers **F-Z)** with no significant differences between groups. **V&X)** Eotaxin-3 and MDC (CCL22) were separated by sex; and **Z)** by treatment (T) with or without (UT) L-dopa; due to a priori identification of sex and treatment difference, respectively. Twenty-six extra/additional HCs serum samples from Exp#1 were added for biomarkers unaffected by extra freezing/thaw cycle **F**, **H, I, L)** more info in **Suppl.Table3**. Lower and upper limit of quantification (LLOQ & ULOQ) are plotted if relevant.

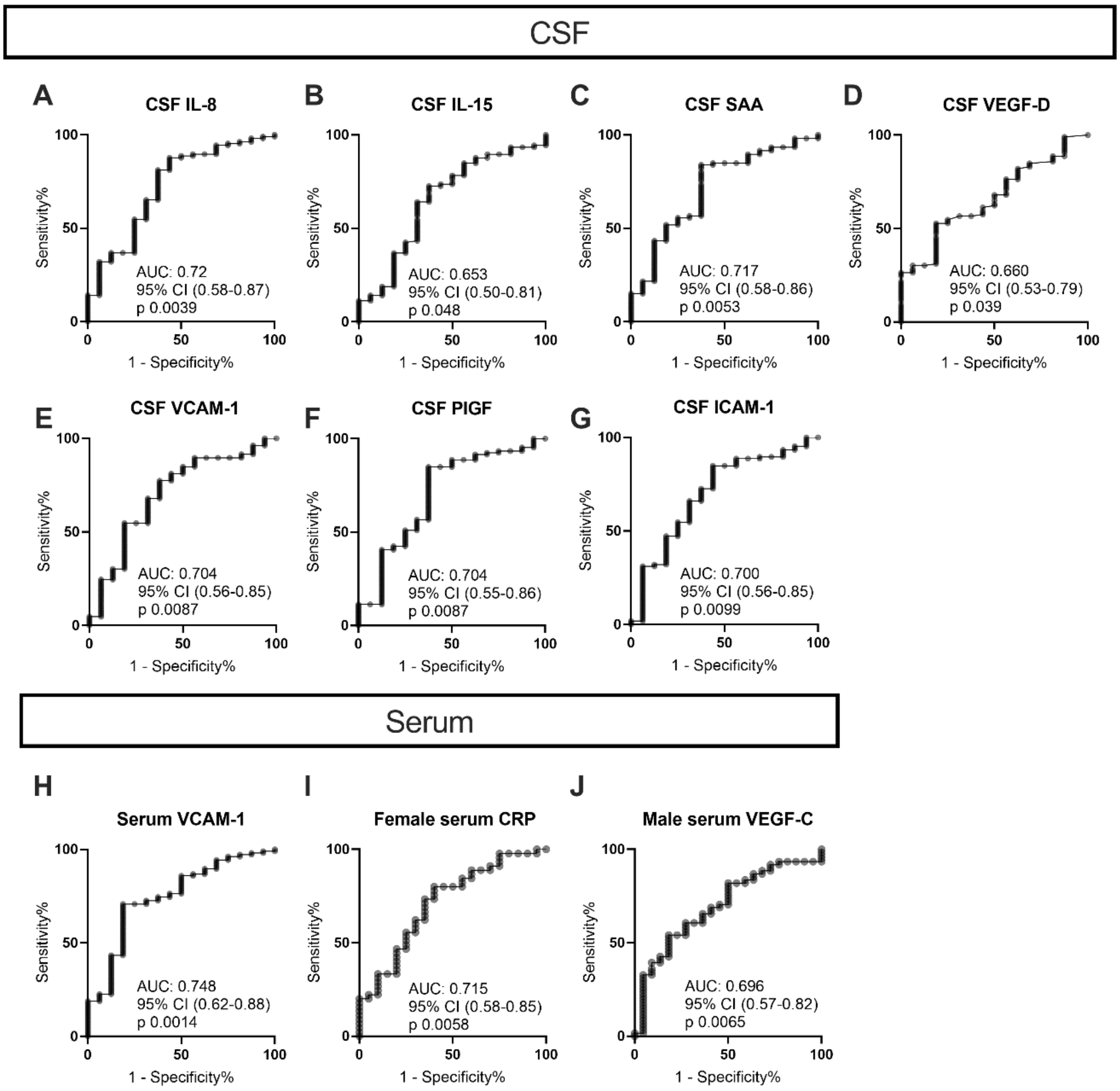

**Suppl. Fig. 3 ROC plots for single biomarkers effect on PD prediction estimated by AUC (Exp#2)**
The ROC curves with p value ≤0.05 for biomarkers ability to predict PD diagnosis in CSF **A-G)** and serum **H-J)** except for sCD163 (see main text **Fig.5**). **A-H)** no sex-discrepancy, **I)** CRP for females only, **J)** VEGF-C for males only. Area under the ROC curve (AUC), confidence interval (CI).

**A**

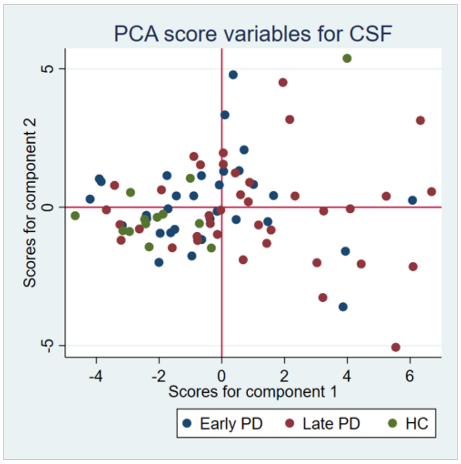

**B**

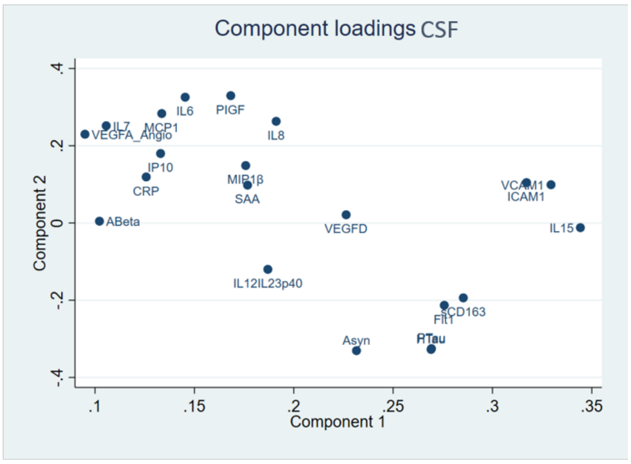

**C**

| Variable | Comp1 | Comp2 |
|---|---|---|
| sCD163 | 0.2853 | -0.1939 |
| ICAM1 | 0.3172 | 0.1046 |
| IL15 | 0.3443 | -0.0121 |
| VCAM1 | 0.3295 | 0.0990 |
| Flt1 | 0.2758 | -0.2131 |
| IL12IL23p40 | 0.1870 | -0.1200 |
| IP10 | 0.1330 | 0.1799 |
| IL8 | 0.1912 | 0.2633 |
| PIGF | 0.1683 | 0.3297 |
| MIP1β | 0.1758 | 0.1487 |
| MCP1 | 0.1336 | 0.2834 |
| VEGFD | 0.2264 | 0.0213 |
| CRP | 0.1258 | 0.1192 |
| IL6 | 0.1454 | 0.3258 |
| IL7 | 0.1056 | 0.2517 |
| SAA | 0.1767 | 0.0981 |
| VEGFA_Angio | 0.0949 | 0.2298 |
| ABeta | 0.1022 | 0.0047 |
| HTau | 0.2691 | -0.3274 |
| PTau | 0.2693 | -0.3253 |
| Asyn | 0.2316 | -0.3305 |

**Suppl. Fig. 4 Principal component analyses for CSF biomarkers effect on PD prediction (Exp#2)**
Principal component analysis (PCA) was computed for all reliable biomarkers in CSF for Exp#2 for separation of the healthy control (HC) group and the early and late Parkinsons disease (PD) groups. **A)** Variables with standardize scales are reoriented on a two dimensional plot of two new principal component variables based on covariance matrix computed eigenverctors. Each dot represents a study participant including 13 out of 16 HC and 70 out of 106 PD patients from Exp#2. **B)** Plot of component loading behind the PCA score. **C)** Variables (CSF biomarkers) effect on component 1 and 2.

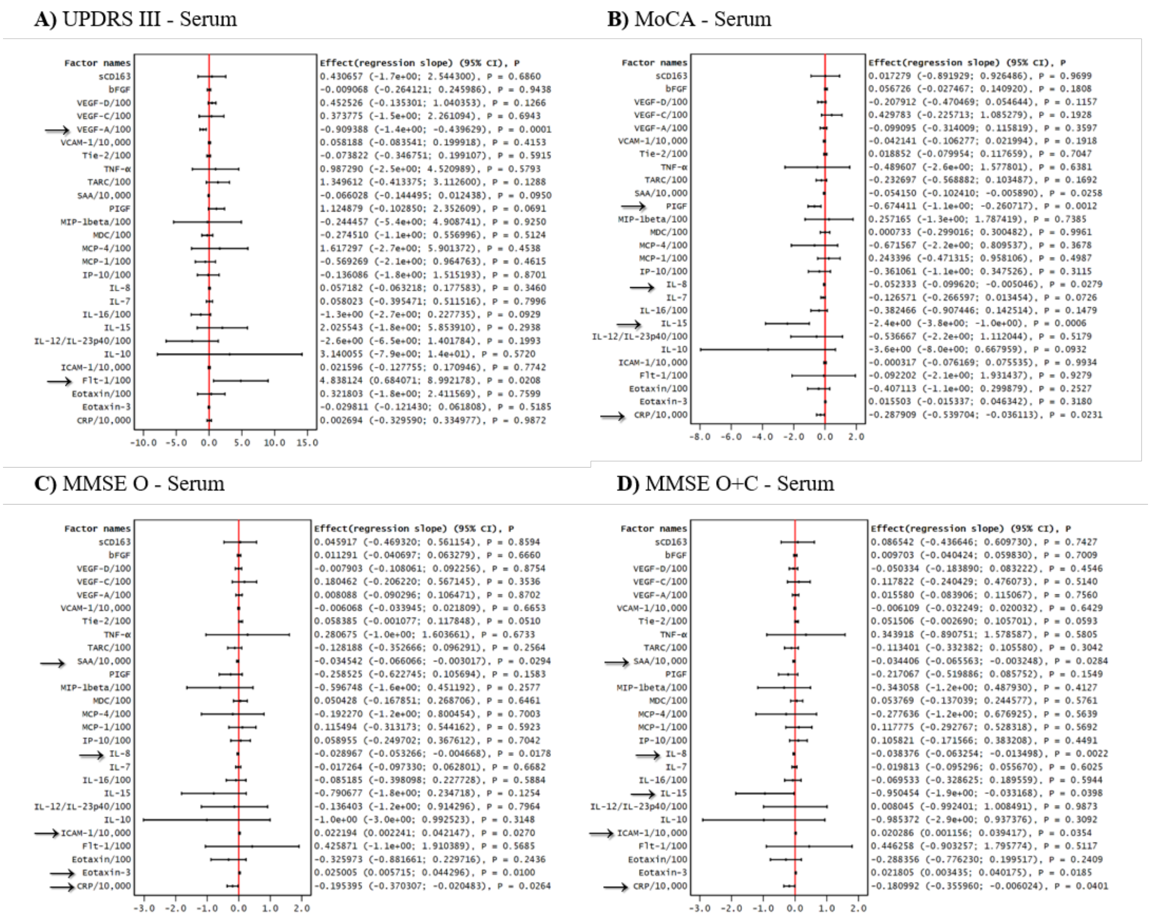

**Suppl. Fig. 5 Serum, Forest plots: Main effect of single biomarkers (Exp#2)**

Predicting effect of single serum biomarkers as forest plots with respect to phenotypical scores by **A)** UPDRS III, **B)** MoCA, **C)** Mini Mental (MMSE original (O)), and **D)** MMSE original or converted from MoCA (O+C). All analytes are measured in pg/mL, except sCD163: mg/L. Some variables are re-scaled (stated in plot) for better presentation. Variables with significant p value are marked with an arrow.

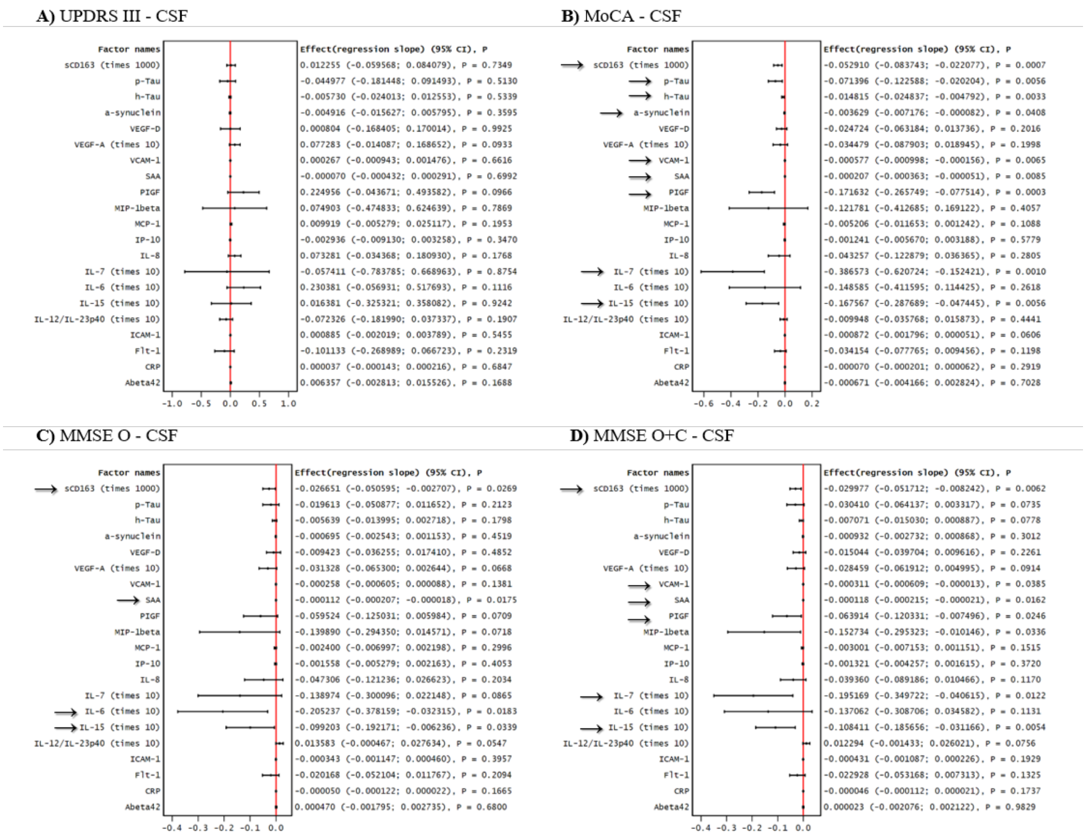

**Suppl. Fig. 6 CSF, Forest plots: Main effect of single biomarkers (Exp#2)**
Predicting effect of single CSF biomarkers as forest plots with respect to phenotypical scores by **A)** UPDRS III, **B)** MoCA, **C)** Mini Mental (MMSE original (O)), and **D)** MMSE original or converted from MoCA (O+C). Variable sCD163 is measured in mg/L and is re-scaled with 1000 to get a better presentation. All other variables are measured in pg/mL with IL-6, IL-7, IL-15, IL-12/IL-23p40, and VEGF-A being re-scaled by 10. Variables with significant p value are marked with an arrow.

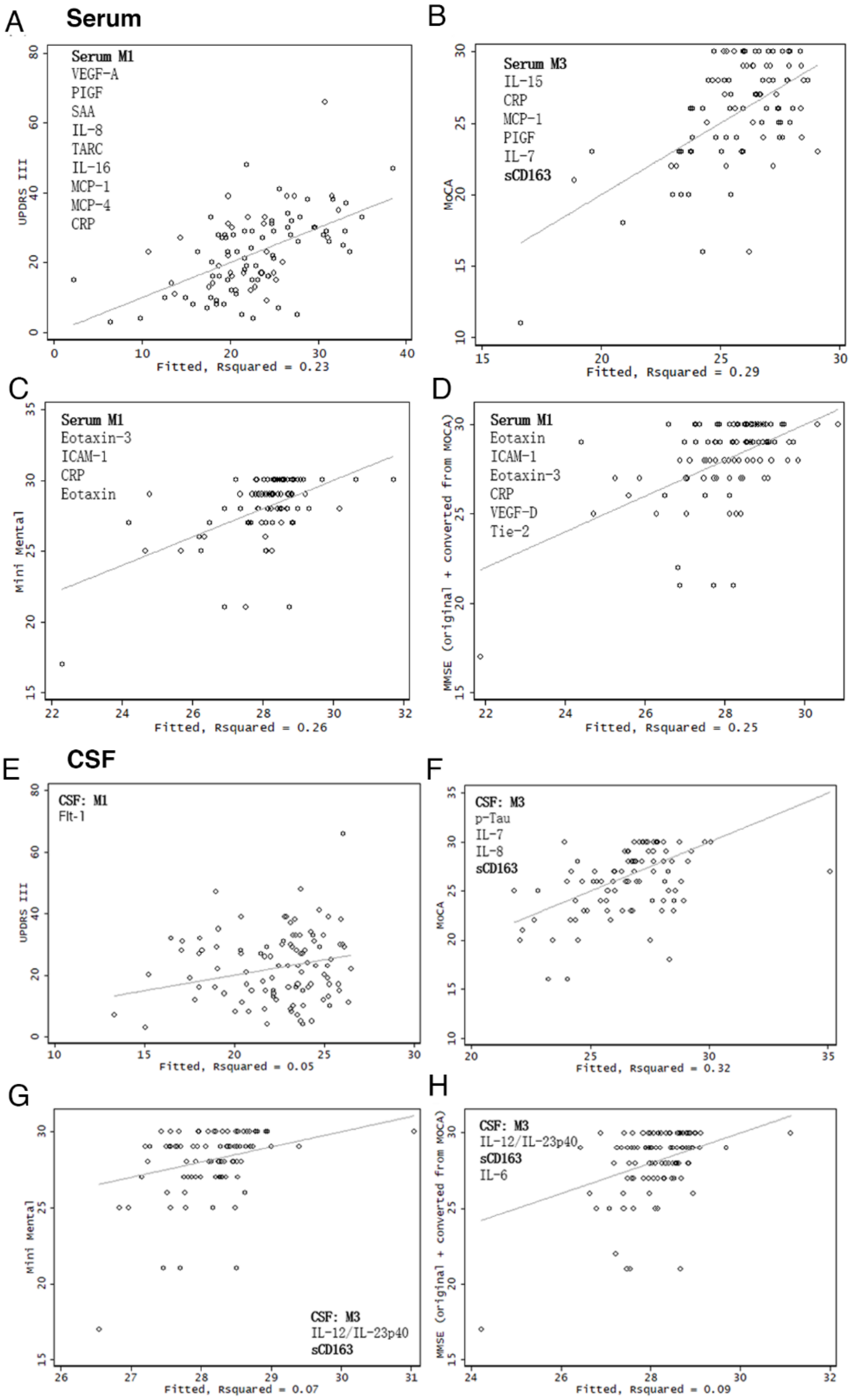

**Suppl. Fig. 7 Serum and CSF the goodness of fit from selected regression models (Exp#2)**
Stepwise forward model strategies (**M1:** probability of entry is 0.1, probability of removal is 0.2; **M2:** probability of entry is 0.05, probability of removal is 0.1; **M3:** probability of entry is 0.1, probability of removal is 0.2, CD163 forced to be included) were used for a linear regression with robust variance component estimation for serum **A-D)** and CSF **G-H)** biomarkers with effect on PD progression scores: **A&E)** Unified Parkinson's Disease Rating Scale three (UPDRS III), **B&F)** Montreal Cognitive Assessment (MoCA), **C&G)** Mini Mental original score (MMSE O), or **D&H)** MMSE original score plus converted from MoCA (MMSE O+C). The models are compared using AIC and BIC and goodness of fit (R squared) **(Suppl.Table 6&7)**. The best models for each prediction score are plotted as fitted values versus measured values.

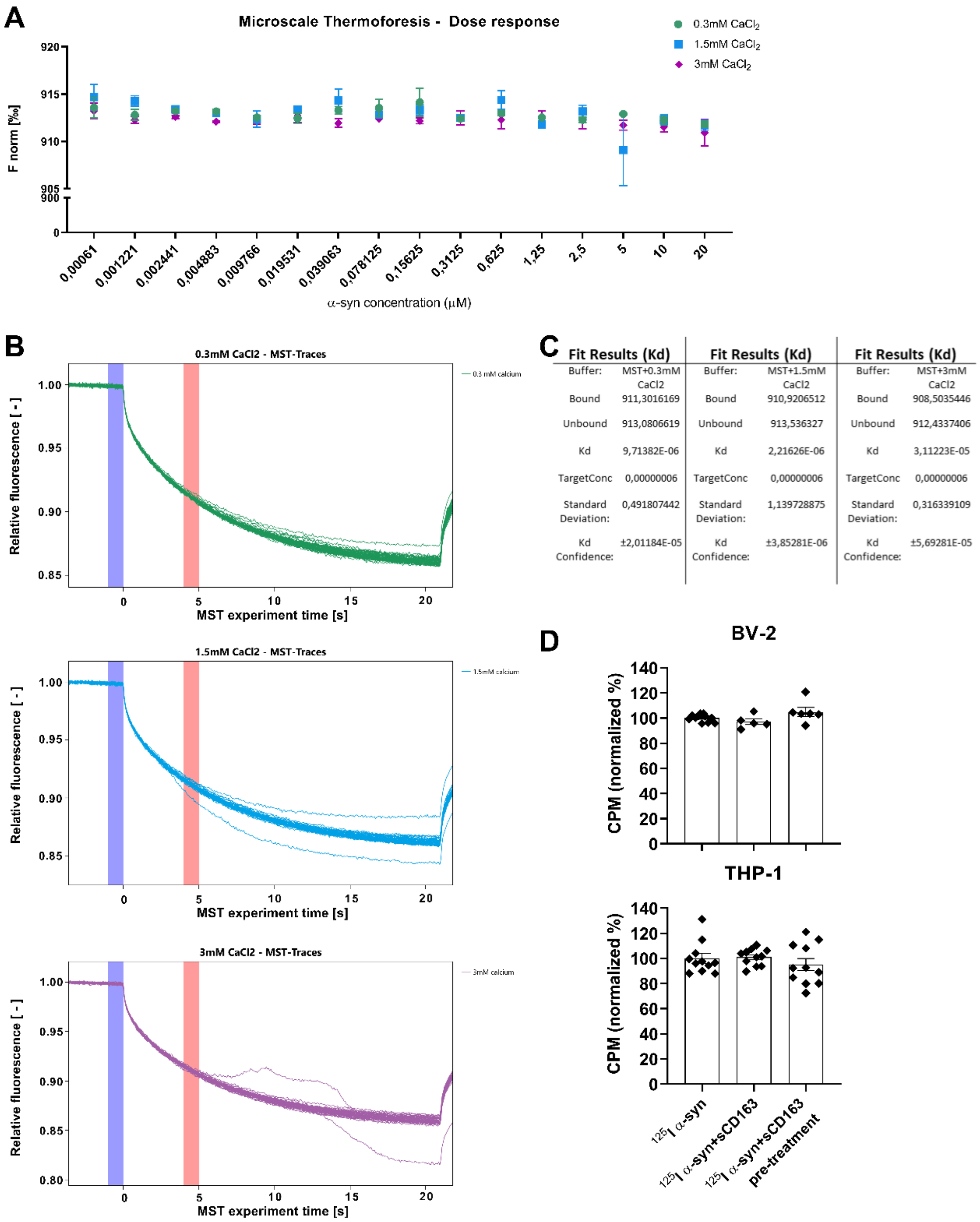

**Suppl. Fig. 8 No direct binding between sCD163 and human α-syn *in vitro***

The interaction between α-syn and sCD163 was studied by Microscale Thermophoresis (MST). Fluorescently labeled (REDtris-NTA-647) human sCD163 was titrated by recombinant unlabeled human α-syn at various concentrations. **A)** Calcium-dependent binding was tested by adding increasing $CaCl_2$ concentrations to MST buffer. **B**) MST traces for each $CaCl_2$ concentration: 0.3mM, 1.5mM, 3mM. Blue- and red-shaded vertical columns represent Fcold and Fhot regions, respectively, used to calculate Normalized Fluorescence (F norm). No binding was detected in any condition. **C)** The curve and Kd values were calculated by averaging Kd curves assimilated using NT analysis software from three independent experiments. **D**) BV-2 and differentiated THP-1 cells were incubated at three different conditions: With iodine-labeled α-synuclein ($^{125}$I α-syn) pre-formed fibrils (PFF) (control); co-incubated with $^{125}$I α-syn and sCD163 (5μg/mL); or pre-treated with sCD163 (5μg/mL) 30 min prior $^{125}$I α-syn addition to the media. Cells were incubated at 4°C (1h) to evaluate and confirm no differences in surface binding. Radioactive counts per minute (CPM) on THP-1and BV-2 were normalized to α-syn average counts and shown as % of control internalization/binding. Statistics: Two-way ANOVA followed by post-hoc Tukey's multiple comparison test when appropriate. Data is shown as mean ± standard error of mean.
